# Unveiling paths to Alzheimer’s disease: excitation-inhibition ratio shapes hierarchical dynamics

**DOI:** 10.1101/2025.02.25.25322663

**Authors:** Francesca Saviola, Asia Ferrari, Daniele Corbo, Michela Pievani, Silvia Saglia, Annamaria Cattaneo, Ilari D’aprile, Giulia Quattrini, Valentina Cantoni, Enrico Premi, Barbara Borroni, Roberto Gasparotti

## Abstract

Alzheimer’s disease is marked by cognitive and memory impairment, with early disruptions in the balance between excitatory and inhibitory neurotransmission, thought to be closely linked to co-occurrent brain changes. The posterior-to-anterior hypothesis posits that functional neurodegeneration begins in critical areas of the default mode network, particularly the hippocampus and posterior cingulate cortex, before extending to more anterior brain regions. This study seeks to evaluate how cortical hierarchy, measured with functional connectivity proxies for excitation/inhibition equilibrium, shapes across the continuum from cognitively unimpaired individuals to symptomatic Alzheimer’s disease.

We include 97 participants: 28 patients (including 20 carriers of the Apolipoprotein E allele, ɛ4+), 35 at-risk individuals (ɛ4+), and 34 controls (ɛ4-). Resting-state functional MRI and T1-weighted imaging were collected for all subjects, with a subset of Alzheimer’s patients also undergoing GABA-edited magnetic resonance spectroscopy in posterior cingulate cortex to provide a multimodal description of pathology-related excitation/inhibition disruptions’ impact on cortical hierarchical dynamics. To probe the validity of excitation/inhibition proxies, we (i) investigated the relationship between *in-vivo* measurements and cognitive profile in Alzheimer’s; (ii) compared default mode network temporal dynamics across groups; (iii) tested its multivariate association with cognitive profile and genetic interactions; (iv) and quantified subject fingerprints related to both pathology presence and genetic risk factors.

The *in-vivo* excitation/inhibition ratio significantly related to cognitive deficits in Alzheimer’s patients, indicating that lower inhibition corresponds to poorer cognitive performance. A voxel-wise analysis demonstrated a positive association between neurometabolism in the posterior cingulate and temporal dynamics across default mode network regions, which can effectively differentiate between patients and controls. Furthermore, network fluctuations showed significant links to cognitive performance metrics, particularly among at-risk individuals. The study identified distinct functional fingerprints based on cortical temporal dynamics, emphasizing the interplay between genetic predisposition and the presence of Alzheimer’s disease.

This investigation provides compelling evidence for the clinical importance of functional connectivity proxies related to excitation/inhibition, particularly within the default mode network. Neurodegeneration induces both a temporal and neurometabolic functional regression in higher-order cortical areas, resulting in a loss of specialized function. Consequently, the hierarchical continuum of cortical functions is disrupted, leading to a homogenization of brain activity. Excitation/inhibition proxies can expand our ability to recognize brain fingerprints of at-risk pre-symptomatic and pre-clinical subjects, opening pathways for potential disease-modifying treatments.

## Introduction

Alzheimer’s disease (AD) is a neurodegenerative condition characterized by progressive memory and cognitive decline that ultimately leads to behavioral and functional impairment. The equilibrium between excitatory and inhibitory neurotransmission, regulated respectively by glutamatergic and GABAergic brain transmitters, is essential for maintaining optimal neural circuit activity and sustaining cognition.^1,2^ Disturbances in excitation-inhibition balance (EIB) can emerge as an early indicator of AD and are significantly associated with structural brain alterations, functional networks activity and cognitive decline.^3,4^

Research demonstrated that various brain neurometabolites, mapped *in-vivo* through Magnetic Resonance Spectroscopy (MRS),^5,6^ undergo significant changes with aging. Disruptions in these metabolites and their interrelationships may increase susceptibility to neurodegenerative diseases, such as EIB alterations associated with early AD pathology.^7^ However, the link between EIB dynamics and temporal fluctuations in BOLD signals, especially in regions that are among the first to be impacted by neurodegeneration, remains largely unexplored.

Additionally, the potential of this relationship to serve as an early biomarker for preclinical individuals with only genetic risk factors - prior to the onset of behavioral deficits - has yet to be fully investigated. The major susceptibility gene for AD, the Apolipoprotein E (APOE) ε4 allele, can identify at-risk populations and influence the EIB gradient since preclinical stages.^8^ Recent findings in cognitively unimpaired individuals indicate that whole-brain EIB is negatively correlated with APOE-ε4 carriage (ε4+) and reveal significant variations, to a greater extent than amyloid accumulation, across critical brain regions (e.g. hippocampus and amygdala).^8,9^

From a connectomic perspective, one of the main functional networks involved in AD is the DMN.^10,11^ The DMN is a well-known functional brain network active during rest and internal cognitive processes involving interconnected regions encompassing the hippocampus, retrosplenial and posterior cingulate cortex (PCC), and the medial and dorsal frontal cortex.^12,13^ The trajectory of AD is believed to originate within disease-specific hubs of DMN. Specifically, while tau pathology predominantly affects temporal regions (the entorhinal and hippocampal regions),^14^ amyloid-related cortical alterations are noted in posterior parietal DMN hubs,^15^ highlighting the intricate interconnection of these pathological features in the advancement of AD. Significantly, tau neurofibrillary tangles are closely associated with atrophy^16,17^ and connectivity changes within the posterior DMN,^18,19^ while frontal regions display a remarkable resilience to neurodegeneration despite elevated levels of amyloid-beta.^20^ It is consequently hypothesized that disruptions in functional connectivity progress from posterior to anterior regions (Hampel et al., 2021), along the core functional network,^21^ mirroring prion-like spreading of amyloid and tau brain aggregates.

Beyond functional connectivity measures, surrogate metrics capturing the dynamic interplay between excitatory and inhibitory neural processes can also be extracted from the BOLD signal,^22^ potentially offering insights into AD pathophysiology. A prominent example is Intrinsic Neural Timescale (INT), quantifying the duration for which neural activity patterns persist over time.^23^ These measures are organized along gradients within the brain’s functional architecture, where they exhibit variations based on the biological properties of the cortex. Specifically, they create a continuum ranging from unimodal perceptual regions to transmodal associative areas. Previous research has demonstrated a significant correlation between EIB measures and variance in temporal structure-function coupling, the latter being lower in perceptual regions.^24–26^ This relationship reflects a gradient of EIB balance throughout the brain which is also strictly linked to structure, cognition and pathology.^27–30^

To date, the extent to which EIB proxies can improve our understanding of a subject’s unique functional brain architecture - and how this connects to their identifiability within the continuum of AD pathology - remains unclear. Unlike standard functional connectivity, which may present a broad perspective on brain interactions, EIB proxies could provide more profound insights into the specific patterns of functional organization that define individual subjects. Herein, functional connectome fingerprinting^31^ offers a powerful approach to investigate early alterations in brain networks by identifying individuals and pinpointing specific brain regions that most effectively distinguish those with particular clinical or genetic conditions from control groups.

In this work, we explore the hypothesis that EIB proxy measures are sensitive to early neural modifications in cognitively unimpaired subjects at-risk for AD, making it a potential biomarker for preclinical AD. Our first aim is to investigate EIB proxy measures gradients from healthy controls, via genetically at-risk individuals, to symptomatic AD patients. The second aim explores associations between *in-vivo* MRS-derived EIB measurements and EIB proxies through functional neuroimaging in disease-specific hubs to demonstrate its biological relevance. Furthermore, by employing functional connectome fingerprinting informed with INT, we aim to assess whether individuals at genetic risk exhibit distinctive patterns from controls that may help disentangle potential progression towards AD. Lastly, we aim at investigating the relationships between EIB, neural fluctuations and cognitive performance to identify markers sensitive to disease progression.

By assessing the dynamics of EIB in the context of AD, we intend to contribute to the identification of early biomarkers and therapeutic targets that could mitigate the progression of this debilitating condition.

## Materials and methods

### Participants

In this study, two separate cohorts of participants were included. Specifically for the first sample, 28 AD patients according to current clinical criteria,^32^ including 20 carriers of the Apolipoprotein E ((APOE) ɛ4 allele, ɛ4+), were recruited at the Neurology Unit, University of Brescia, Italy. Exclusion criteria include cerebrovascular disorders, hydrocephalus, intracranial masses documented by MRI, a history of traumatic brain injury, and/or serious medical illness.^33^ The second sample included 69 cognitively unimpaired (CU) older adults recruited at the IRCCS Istituto Centro San Giovanni di Dio Fatebenefratelli in Brescia, Italy.^34^ This cohort comprised 35 carriers (ɛ4+) and 34 non-carriers (ɛ4-) of the APOE ɛ4 allele. Age≥60 years and the capacity to give written informed consent were the inclusion requirements. Exclusion criteria were a MMSE≤24, pathological results on at least two standardized cognitive tests (Supplementary material), being carrier of an autosomal dominant genetic mutation (APP, PSEN1, PSEN2), the presence of depressive symptoms or major psychiatric or neurological conditions.^34^

### MR Acquisition

Functional MRI (fMRI) data were acquired for both AD and CU groups, while Magnetic Resonance Spectroscopy (MRS) was performed only on a subsample of the AD group. The complete MRI protocol, including detailed acquisition parameters for both groups, comprehensive preprocessing steps, and quality control procedures for fMRI and MRS data, are fully described in Supplementary materials.

### Intrinsic Neural Timescale (INT) estimation

Using the anatomical Glasser atlas,^35^ 360 cortical parcels were created from the preprocessed rs-fMRI data. The obtained matrices were then employed as inputs for a model-free estimation of intrinsic timescale, where we calculated lagged (auto-) covariance using blocks of contiguous time points, to uncover the dynamic fluctuations within each parcel. To maintain consistency in time delays across varying temporal resolutions (TR), for the covariance lags initially 6 were employed for the first group (Lags_AD-group_), reflecting the similarity in TR with the original study.^23^ For subsequent groups with different TRs, appropriate number of lags were calculated as follow: Lags_AD-group_*TR=Lags_APOE-group_. Through the computation of the zeros of a spline fit to the autocovariance function (ACF) (https://github.com/ryraut/intrinsic-timescales),^23^ the precise abscissa corresponding to an autocorrelation value of 0.5 (i.e half of the ACF full width at half maximum) was estimated (see Raut et al.,^36^ for extensive procedure). Notably, a slower decay in ACF is often interpreted as indicative of longer intrinsic timescales, suggesting that the brain’s neural responses take more time to return to baseline after a stimulus, thereby reflecting sustained neural activity patterns and prolonged engagement of neural processes. ACF values and INT were calculated based on our a priori hypothesis, focusing exclusively on parcels located within the hippocampus or regions corresponding to the DMN functional network (7 networks parcellation)^37^ and within the anterior and posterior parts of the latter (Supplementary Fig. 4).

### Functional fingerprinting informed by INT

To calculate the functional connectivity (FC) matrices of each participant we parcellated the 4D fMRI images with the same atlas employed for the INT estimation.^35^ In order to investigate individual functional connectivity identifiability informed by INT according to disease and genotype, for every subject FC matrices computed on the first half of the rs-fMRI time course (half volumes) as test, and FC matrices computed on the second half of the rs-fMRI time course as retest.^31^ Subsequently, Pearson’s correlation coefficients between time series from all atlas regions were computed to create a 360 x 360 FC matrix for each subject and time point and multiplied for corresponding INT vector to informed FC weights.

To assess FC profiles according to APOE genotype (ε4+ vs. ε4-) and cognitive status (AD vs CU), we created an identifiability matrix^31^ where rows represent INT-weighted FCs from the test session and columns represent those from the retest session. We calculated Pearson’s correlation coefficient to measure similarity. The average of the diagonal elements, termed “self-identifiability” or “*Iself*”, reflects the similarity of a subject’s FC profile across sessions. The average of the off-diagonal elements, known as “*Iothers*”, indicates similarity across different subjects. Based on the functional connectome fingerprint hypothesis, we calculated “differential identifiability” or “*Idiff*” (*Idiff* = (*Iself* - *Iothers*) * 100) to quantify how distinct an individual’s FC profile is within the group.^31^ Furthermore, success rate (SR) was computed as the percentage of subjects correctly identified, meaning those whose test data was correctly matched with their corresponding retest data, out of the total number of subjects. We also examined whether identifiability according to APOE genotype was influenced by specific connectome edges or networks, particularly if certain edges enhanced identifiability in the APOE e4+ group. To analyze this, we computed the intraclass correlation coefficient,(ICC_1,1_)^37,38^ which measures agreement across groups, with higher values indicating stronger agreement. For these analyses, we took the classification of ‘good agreement’ (*ICC* > 0.6) as a reference to threshold the ICC matrices.^39^ Edge-wise ICC was calculated for all potential edges within each group to quantify the functional connectivity fingerprint unique to each group of interest. Subsequently, pairwise comparisons between groups were conducted by categorizing the functional networks into unimodal and transmodal groups, utilizing the Wilcoxon rank-sum test for analysis. The resulting p-values were then adjusted through the application of false discovery rate (FDR) correction to account for multiple comparisons. The matrices were subsequently binarized for ICC values > 0.6 (Supplementary Fig. 5). We aimed to identify the commonalities in the distribution of edges with the highest ICC, both within and between functional resting-state networks. ^25,26,40^ To accomplish this, we followed a previously described procedure^41^ to analyze the ICC binary matrix and computed several metrics. For each of the seven networks, we quantified the number of overlapping ICC edges (ICC_0_) both within and between networks. We then calculated the proportion of ICC0 edges relative to the total number of edges in each network, defining this as pICC_0_. Finally, to assess the distance from the healthy reference group (i.e., CU-ε4-), we computed a ratio based on healthy individuals. This ratio (*Rnet*) was determined by dividing pICC_0_ for each group of interest (e.g., AD-ε4+, AD-ε4-, or CU-ε4+) by pICC0 for the healthy reference and subtracting 1.

### Biological relevance of INT

To test the validity of INT as a proxy measure of EIB balance, we explored its association with direct EIB measures derived from MRS data collected in the AD sample. Therefore, after computing and concatenating the single-subject whole-brain INT maps, resulting images were taken to group-level analysis using FSL’s randomise.^42^ We performed voxelwise nonparametric inferential statistics (TFCE, 5000 permutations) to identify clusters of INT patterns associated with the neurometabolites of interest (GABA+, Glx, and EIB balance). We examined whole-brain INT patterns through a GLM using additional covariate (i.e., gender as a covariate of no interest and neurometabolites as a covariate of interest). FWE correction across voxels (*p* < 0.05) and Bonferroni correction across tests (*p* < 0.02) were applied. Peak activations were then matched with the chosen parcellation scheme^35^ to pinpoint the specific anatomical regions.

### Brain-Behaviour relationship

Partial Least Squares Correlation (PLS-C) methodology was employed to disentangle the complex relationships between cognitive and brain imaging features. This sophisticated multivariate approach identifies latent variables that optimize the correlation between two multidimensional datasets. In our study, these datasets comprised DMN INT values and an array of cognitive performance measures. The analysis included 97 subjects, incorporating 101 DMN parcels and 11 behavioral variables. The latter included ε4 status, scores from various cognitive assessments, and the interactions between genetic predisposition and performance. The comprehensive methodology, encompassing matrix computations, rigorous statistical procedures, and nuanced interpretation of results, is detailed in Supplementary material.

### Statistical analysis

To determine differences in sociodemographic and cognitive variables among the groups, we conducted a series of statistical analyses. Prior to any statistical test, the Shapiro-Wilk test for normality was performed. We evaluated MR quality criteria and demographic variables such as age, education level, and gender distribution using Chi-squared tests for categorical variables and Kruskal-Wallis for continuous variables. For cognitive performance measures, we employed non-parametric Kruskal-Wallis tests to compare MMSE, Rey Auditory Verbal Learning Test and TMT scores across groups. Post-hoc analyses were performed using the Mann Whitney U test with FDR correction to determine specific group differences.

To test associations between imaging and clinical scores, statistical analysis was conducted using advanced modeling techniques. For the MRS sample, Generalized Additive Models for Location Scale and Shape (GAMLSS) with a zero-adjusted Gamma distribution (ZAGA) were used, suitable for non-normal data with zeros. Regarding MRS-derived metrics (Glx, GABA+ and EIB), in the AD group only, we tested the relationship between the former and clinical scores (i.e. CDR and ADAS-Cog-13).

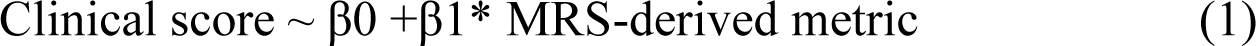

For each clinical assessment, Clinical score represents the score obtained in the test. MRS-derived metric is the neurometabolite of interest.

For the whole sample, a generalized linear mixed model (GLMM) was employed after model comparison. The GLMM used log-transformed INT as the dependent variable, with fixed effects for sex, clinical scores, and their interactions, plus a random intercept for participants. FDR correction was applied for multiple comparisons. Given the association between INT and EIB balance, we repeated these analyses within the DMN, further dividing it into anterior and posterior regions. To explore the interaction between cognitive impairment (AD/CU) and APOE status (ε4+/ε4-), and to be consistent with the PLS-C analysis,^43^ we transformed the relevant variables into z-scores and created an interaction variable by multiplying these z-scores. This interaction term captures the combined effect of cognitive and APOE status on the dependent variable, enabling us to incorporate both information in the same model.

We tested INT within the DMN to compare the four groups (AD-ε4+, AD-ε4-, CU-ε4+, and CU-ε4-) while controlling for gender and accounting for individual variability through random effects. To achieve this, the log-normal model was identified as the most suitable for capturing the underlying relationships while effectively addressing issues of overdispersion and non-normality in the dataset (Supplementary material).

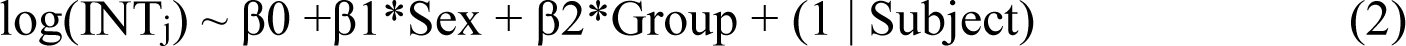

For each region j belonging to the DMN, INT_j_ is its INT score. Sex is the term controlling for gender effect, whereas Group is a factor (i.e. 4 levels, AD-ε4+, AD-ε4-, CU-ε4+, CU-ε4-).

Then, we assessed the relationship between INT within the DMN and MMSE across the four groups (AD-ε4+, AD-ε4-, CU-ε4+, CU-ε4-).

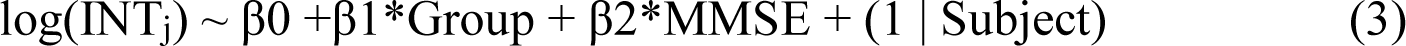

Both models were performed on anterior and posterior DMN separately as well as within the hippocampus.

For brain fingerprinting analysis, we initially assessed the success rate (SR) for all groups. Following this, we conducted Kruskal-Wallis tests to compare the metrics (i.e., *Idiff*, *Iself*, and *Iothers*) and examine differences among the various groups (i.e. AD-ε4+, AD-ε4-, CU-ε4+, CU-ε4-). Lastly, we sought to determine whether there was a significant difference in the distribution of significant edges for identification across groups for each of the functional networks. To accomplish this, we performed a Wilcoxon test for each network, comparing the number of ‘good’ ICC edges (*ICC* > 0.6) in the networks across groups. Significance was adjusted using FDR correction for multiple comparisons accounting for the number of comparisons (i.e., *n* = 28: 7 functional networks and 4 groups).

### Data availability

Data supporting the findings of this study are openly accessible. The analysis code is available on GitHub: https://github.com/ferrariasia06/PATHFIND. For raw material requests related to the Alzheimer’s disease cohort (from BrainSync-AD)^33^, please contact. Requests concerning raw materials from the cognitively unimpaired subjects cohort (from NEST4AD)^34^ should be directed to.

## Results

### Clinical and cognitive profile of the sample

While age, education, and gender were similar across groups (*p* > 0.05), cognitive performance differed significantly between AD patients and both CU-ε4+ and CU-ε4-groups for all tests (*p* < 0.001). Detailed clinical and cognitive scores for the AD-ɛ4+ and AD-ɛ4-subgroups are reported in the Supplementary results.

### *In-vivo* detected EIB in AD relates to INT

Voxel placement coherence and the group-averaged edited GABA+ and Glx spectra from the PCC (gray matter fraction: 0.43 ± 0.06%) in AD patients are shown (Supplementary Fig. 1). After MRS quality assurance, 19 out of 20 subjects were retained (Supplementary Table 4). Across the entire AD group, the following neurometabolite concentrations (in i.u.) were observed: GABA+/Water (3.5 ± 0.5), Glx/Water (13.9 ± 2.1), and the corresponding EIB ratio (4.0 ± 0.7) (Fig. 1A).

**Figure 1.**
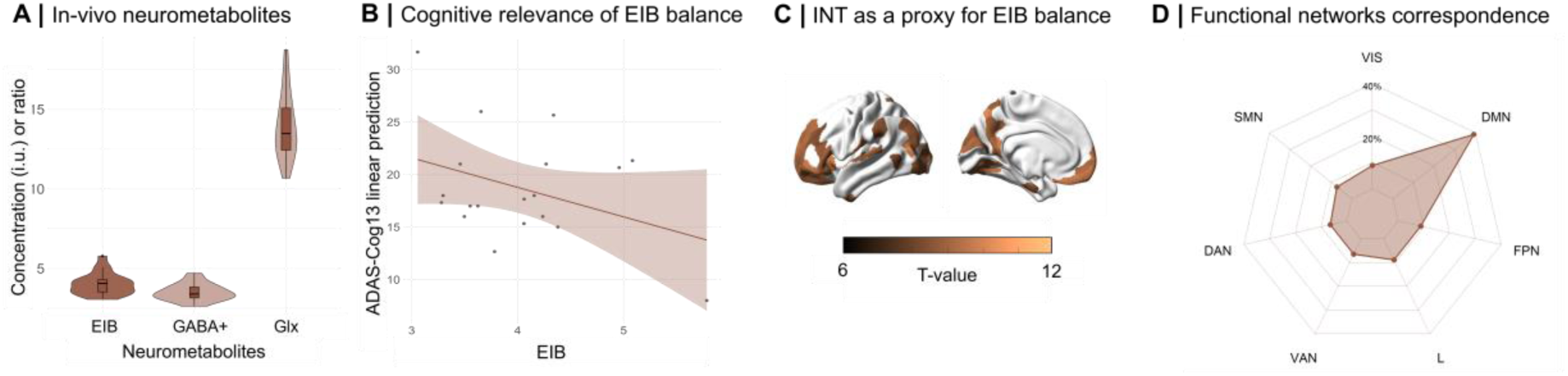
Intrinsic neural timescale (INT) association to neurometabolism in Alzheimer’s disease patients. (**A**) Violin plot of neurometabolite concentrations (Glx and GABA+) and ratio (EIB) measured by MRS in posterior cingulate cortex. (**B**) Linear negative relationship between EIB ratio and cognitive performance. (**C**) Voxel-wise associations between INT and EIB highlighting the strength of correlations between EIB and INT values. (**D**) Functional networks distribution of the voxel-wise associations between INT and EIB. Abbreviations: Glx: Glutamate+Glutamine, EIB: excitation/inhibition balance, VIS: visual network, DMN: default mode network, FPN: fronto-parietal network, L: limbic network, VAN: ventral attention network, DAN: dorsal attention network, SMN: somatosensory network.

Our analysis revealed significant relationships between these neurometabolites, clinical scores and INT in AD patients. Specifically, EIB ratio was negatively associated with ADAS-Cog scores (*β* = -0.25, *p_FDR_* < 0.05; Fig. 1B), highlighting its role in cognitive performance, driven by GABAergic content (Supplementary material). In the AD cohort, INT patterns exhibited a positive voxel-wise relationship with the neurometabolites of interest and the corresponding EIB ratio. Cluster peaks associated with the EIB ratio fell within the frontal medial cortex (voxel-wise TFCE *p_FWE_*< 0.02, Fig. 1C; Table 2), corresponding to the anterior portion of the DMN (Fig. 1C). Notably, the voxel-wise association map encompassed a larger spatial distribution comprehending mainly regions of the DMN (40%; Fig. 1D) and to a lesser extent areas of the other networks (<10%; Fig. 1D). Similar findings were observed for the combined neurometabolites GABA+ and Glx, as detailed in the supplementary materials.

### Intrinsic Neural Timescale

A significant difference was found in whole brain ACF decay between the groups (*χ²* = 9.3, *p* = 0.01; Fig. 2A). Subsequent pairwise comparisons at the whole-brain level indicated that the AD group significantly differed from both the CU-ɛ4+ (*Cohen’s d* = -0.1; *z* = -2.5, *p* = 0.01) and CU ɛ4- (*Cohen’s d* = -0.1; *z* = -3.0, *p* = 0.002) groups, showing a faster decay in the AD group. Specific results related to the DMN are presented in the Supplementary materials There was no significant difference between CU ɛ4+ and CU ɛ4- (*z* = -0.7, *p* > 0.05), underline absent specificity of whole brain ACF decay in detecting differences between at risk subjects and CU.

**Figure 2:**
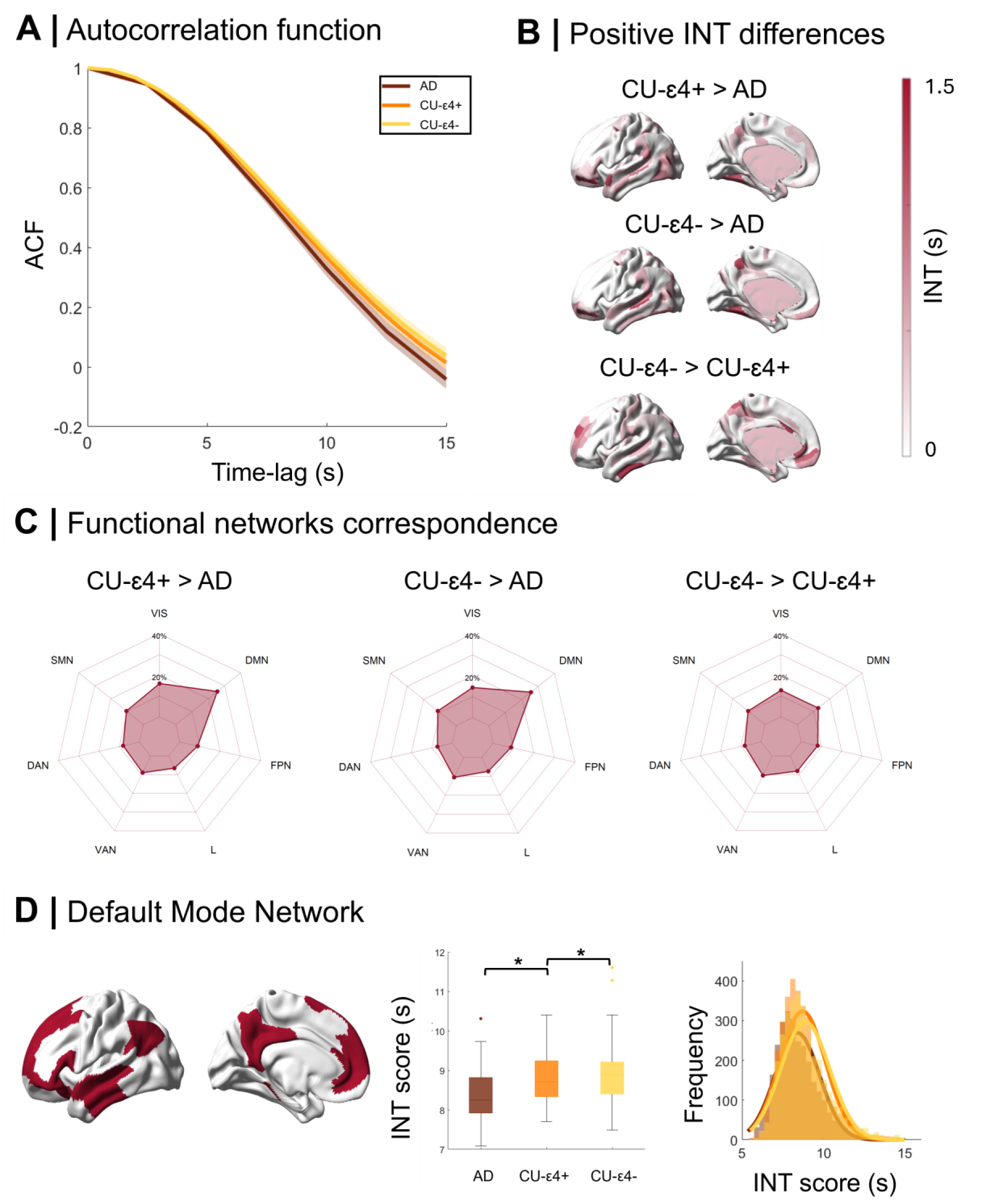
Intrinsic neural timescale (INT) differences between groups. (**A**) Intrinsic timescale was estimated for each cortical parcel as the temporal autocorrelation decay during the resting state, quantified as half of the full width at half maximum of the ACF for each group. (**B**) Cortical positive differences of mean intrinsic timescale across groups pairs. (**C**) Functional networks distribution of differences in INT between AD, CU-ɛ4+ and CU-ɛ4-. (**D**) Left: Brain renders of DMN, Center: Global mean of regional INT in DMN, Right: Distribution of regional INT in DMN.

When testing the differences in INT across groups, a significant difference in the intrinsic neural timescale at the whole-brain level was found between the groups (i.e. AD, CU ɛ4+, CU ɛ4-) (*η2* = 2, *χ²* = 738.4, *p* < 0.0001). Subsequent pairwise comparisons indicated that the AD group showed significantly shorter INT compared to CU-ɛ4+ (*Cohen’s d* = -0.4; *z* = - 21.5, *p* < 0.0001, Fig. 2B) and CU-ɛ4- (*Cohen’s d* = -0.6; *z* = -26.3, *p* < 0.0001; Fig. 2B). Additionally, a significant difference was observed between CU-ɛ4+ and CU-ɛ4-groups (*Cohen’s d* = -0.2; *z* = -6.4, *p* < 0.0001; Fig. 3B). The functional correspondence of these differences to the Yeo functional atlas is reported in the Supplementary materials. Large effect sizes indicate that the alterations in the frequency of INT are substantial, particularly between AD patients and both CU ɛ4 carriers and non-carriers. More precisely, differences in INT were observed across groups (whole DMN: *χ²* = 221.4, anterior DMN: *χ²* = 89.8, posterior DMN: *χ²* = 142.77, hippocampus: *χ²* = 39.37; *p* < 0.0001). Pairwise comparisons showed significant differences between AD-ε4+/AD-ε4-, AD-ε4+/CU-ε4+, AD-ε4+/CU-ε4-, AD-ε4-/CU-ε4+, AD-ε4-/CU-ε4-, and CU-ε4+/CU-ε4-groups across various regions (p<0.05) (Supplementary material).

**Figure 3:**
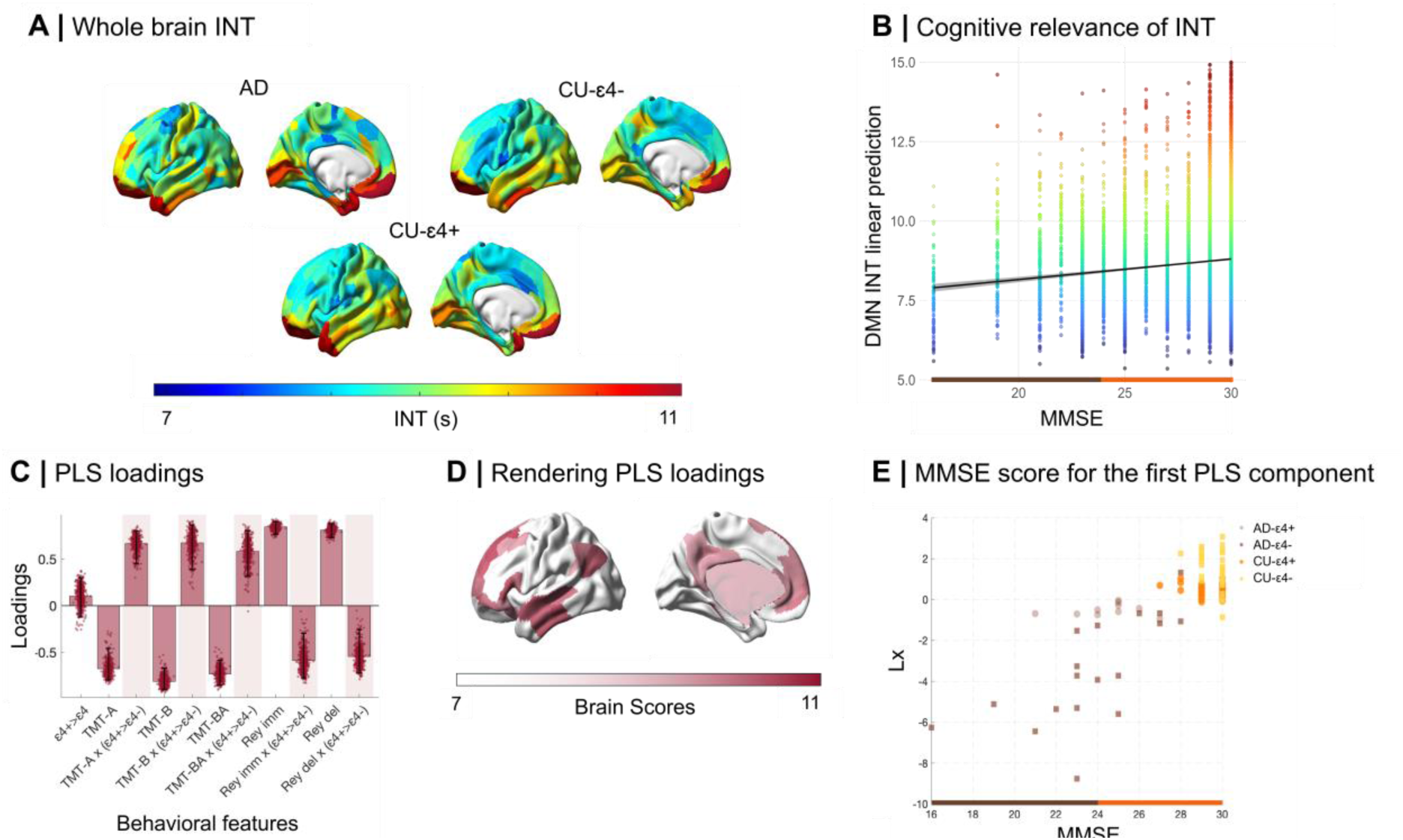
Intrinsic neural timescale (INT) reductions in Alzheimer’s disease. (**A**) Cortical map of mean INT across groups. (**B**) Linear positive relationship between INT within the DMN and Mini-Mental State Examination across groups. (**C**) Significant loadings for the behavioural scores of the detected latent component. (**D**) Significant loadings for the brain scores of the detected latent component projected onto the surface. (**E**) Genetic high risk-score for the latent component colored by group projected on MMSE score.

Linear models, while controlling for head motion, revealed that INT was predicted by Sex (*β* = 0.024, *p_FDR_* = 0.02). and Group (AD-ε4+<CU-ε4-: DMN (*β* = -0.06, *p_FDR_* < 0.01), posterior DMN (*β* = -0.07, *p_FDR_* < 0.01), anterior DMN (*β* = -0.05, *p_FDR_* < 0.01)). Furthermore, while controlling for head motion, INT was predicted by MMSE (DMN: *β* = 0.006, *p_FDR_* = 0.01; posterior DMN: *β* = 0.008, *p_FDR_* = 0.004, Fig. 3B). When using CU-ε4-as a reference group, we observed significant differences between this group and both AD-ε4+ and AD-ε4-groups, but no significant effects were found in the CU-ε4+ group. Notably, when MMSE and Sex were included in the same model, Sex was no longer significant (Supplementary material), suggesting that MMSE alone better explains the model.

Multivariate analysis, performed with PLS-C yielded one significant latent component (LC). Fig. 3C represents pairs of brain regions belonging to the DMN and behavioural variables contributing the most to the multivariate correlation. The LC (*p* < 0.001) highlights a mainly positive significant relationship between INT loadings in DMN regions and all behavioural variables interacting with ε4 carrier variables. The interaction terms between behavioral variables and presence of risk allele seemed to dominate this component (95% variance explained) with higher behavioural saliences for all variables. To better interpret each subject’s contribution in the brain-cognition correlation patterns and how it differs in the presence of ε4 carriage, we plotted the computed interaction for ε4 carrier score projected on the MMSE of subjects (Fig. 3E). Subjects grouped in linear patterns depending on the presence of symptoms and/or ε4 carrier genetic risk factor.

In the fingerprinting analysis, we found that the SR at which subjects were identified based on their FC informed with INT was equal to 100% for all groups. When comparing observed SR and Idiff computed on the single identifiability matrices against their correspondent null distributions (see Methods)^44,45^, a statistically significant effect was obtained (permutation testing, *p* < 0.001) for each group (see Supplementary material). More specifically, group Idiff comparisons (Fig. 4B; *χ²* = 14.6, *p* < 0.01) revealed that Idiff in AD-ε4-was significantly lower than Idiff in CU-ε4+ (*p_FDR_* < 0.05, Wilcoxon test, Table 3). Notably, no significant differences were found between AD-ε4+ and CU-ε4+. To better explore what drove this effect of group in Idiff, we looked at the trajectories of Iself and Iothers distributions separately. We found (Table 3; *χ²* = 37.9, *p* < 0.001) that Iothers was significantly higher in AD-ε4+ than CU-ε4+ and CU-ε4- (*p_FDR_* < 0.05, Table 3). No significant differences were found for Iself.

**Figure 4:**
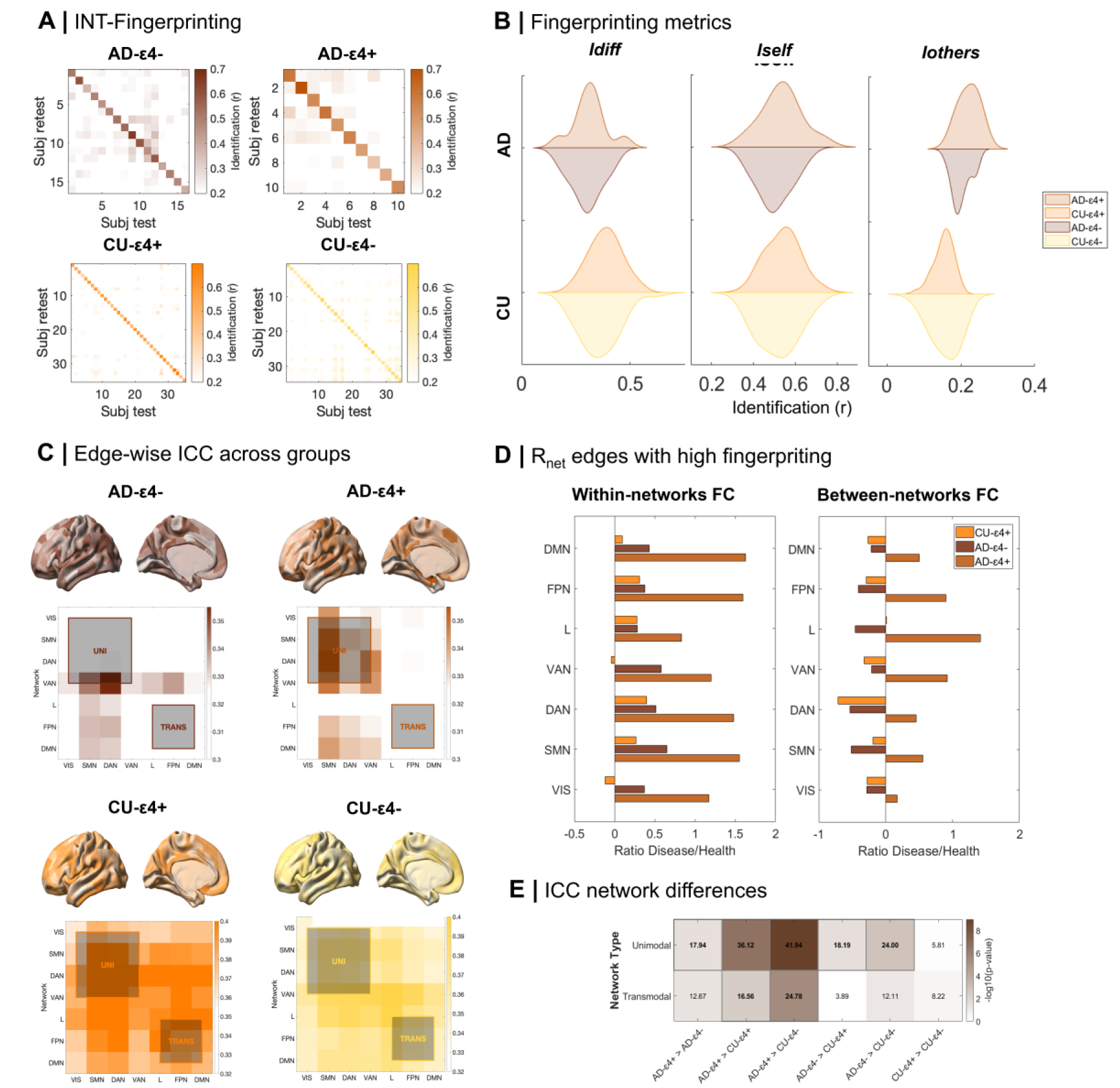
Functional connectome identifiability informed by INT. (**A**) Functional connectome identifiability matrices enhanced by INT. (**B**) Functional connectome metrics compared across groups. (**C**) Surface render of the distribution of ICC (5th to 95th distribution percentile) and average ICCs (>0.3) within and between functional networks across groups. Functional networks have been organized into unimodal/multimodal regions (upper-left square) and transmodal regions (lower-right square). (**D**) Distribution of edges with the highest ICC shared across all groups (*R_net_*), both within networks and between networks. The distance from the healthy reference is calculated as the ratio of disease to health. Positive values indicate an increase in the percentage of edges, while negative values signify a decrease. (**E**) Heatmap illustrating the corrected p-values obtained from pairwise comparisons between four groups (AD-ɛ4+, AD-ɛ4-, CU-ɛ4+, and CU-ɛ4-) across ICC of unimodal and transmodal functional networks. Colorbar represents p-values transformed using the negative logarithm base 10 (-log10), together with differences across groups in each cell.

We then examined the spatial specificity of brain fingerprints by analyzing edgewise intra-class correlation (ICC).^31^ This analysis revealed a spatial reorganization of the most identifiable edges in clinically relevant AD (i.e. AD-ε4+, AD-ε4-) (Fig. 4C). Specifically, edges with the highest ICC values exhibited distinct topological distributions across all groups. Compared to CU-ε4-, we found a slight increase in the number of edges with ‘good’ ICC in CU-ε4+, and a substantial increase in AD dementia, with AD-ε4+ carriers showing the highest number of such edges (Table 3). Based on these findings, we further explored the spatial distribution of edges with good ICC within each network, examining both within- and between-network FC relative to the total edge count (Fig. 4D). The patient groups demonstrated a notable increase in within-network FC compared to controls, with AD-ε4+ showing a substantial 67.5% increase in the number of edges contrasting the 22.5% increase of AD-ε4-. This highlights significant alterations in brain network dynamics. On the other hand, CU-ε4+ subjects showed only 8.5% overall increase, aligning more closely with controls. When assessing between-network connections, a striking pattern emerged. AD-ε4+ exhibited a marked elevation in fingerprint edges (i.e., positive ICC) across all functional networks (e.g. 35%). Conversely, both AD-ε4- and CU-ε4+ groups showed decreases in between-network connections (−18.5% and -14.5%, respectively). This increase was relatively uniform across unimodal and transmodal networks, with unimodal networks showing a slightly more pronounced effect. Notably, patients carrying the genetic risk factor (ε4+) displayed the most substantial changes, underscoring the compounding effect of genetic predisposition on network alterations. These findings emphasize the heightened importance of between-network connections in subjects carrying the APOE ε4 allele, suggesting either a potential compensatory mechanism or a distinctive pattern of network reorganization in response to genetic risk and disease progression.

A comparative analysis of the ‘good’ ICC edges reveals distinct patterns across unimodal and transmodal networks (Fig. 4E). The comparison of AD-ε4+ and AD-ε4-unveils significant disparities in unimodal cortices only with the former showing a higher prevalence of surviving edges (*D_unimodal_* = 18, *p* < 0.05). This dichotomy suggests a differential impact of the ε4 allele within the patients’ group on network stability across brain regions. A similar trend emerges when comparing AD-ε4+ to CU irrespective of the presence of the ε4 allele including transmodal cortices too, where patients exhibit higher level of edges respective to both CU-ε4+ (*D_unimodal_* = 36, *D_transmodal_* = 17; *p* < 0.01) and CU-ε4- (*D_unimodal_* = 42, *D_transmodal_* = 25, *p* < 0.001). Furthermore, AD-ε4-show higher number of edges in unimodal cortices only while compared to CU (CU-ε4+: *D_unimodal_* = 18; *p* < 0.01; CU-ε4-: *D_unimodal_* = 24, *p* < 0.001). Network-wise differences are displayed in the Supplementary material.

## Discussion

The gradient of hierarchical cortical dynamics enables the efficient organization of human brain functions, with primary sensory regions delivering modality-specific information and higher-order regions integrating them with cognitive signals in a transmodal manner. Essential to this hierarchy is the complex interplay between excitatory and inhibitory processes necessary for optimal cognitive functioning. What are the implications when this process does not work properly, as in neurodegenerative diseases? In this study, we present evidence of a disrupted hierarchy in transmodal cortices in AD since preclinical stages, i.e. asymptomatic APOE4 carriers, paving the way for future early detection of AD.

In terms of neural circuit architecture, a hallmark of AD is the disruption of FC within the DMN.^19^ These abnormalities appear even before clinical symptoms and cognitive decline, in the so-called preclinical stages of AD, such as in amyloid-positive individuals^46,47^ or ε4 carriers.^48–50^ This suggests a potential role for FC alterations in early AD detection and supports the concept of network-based functional compensation aimed at preserving cognitive performance.

Based on the posterior-to-anterior hypothesis for AD, we investigate how dorsal EIB, especially in key nodes of the DMN as PCC, correlates with overall clinical severity. This exploration is supported by several previous studies demonstrating a link between excitatory-inhibitory interplay in hippocampal and/or DMN regions and (i) early AD pathology,^51–53^ and both (ii) populations at genetic risk of AD.^8,54^ Here, we highlight how higher EIB values (i.e. characterized by less GABAergic content)^55,56^ are associated with worse cognitive performance and might involve larger cortical areas.^57^ In this context, disturbances in EIB can be viewed as a culmination of micro-level synaptic pathology associated with AD. These metabolic shifts represent both the endpoint of underlying changes and the onset of subsequent network disruptions and cognitive impairment.^7^ Indeed, hierarchical cortical dynamics are significantly influenced by changes in EIB, which can be leveraged to investigate disruptions in AD. A key contribution of this work is demonstrating how EIB proxy measures derived from neurovascular coupling signals, such as BOLD from fMRI, can effectively differentiate between patients and controls, particularly within DMN regions.

Previous studies have shown that functionally hyperconnected regions, especially those within transmodal cortices, exhibit lower EIB proxy values in AD populations.^3,58–60^ Notably, INT - the duration over which a brain region processes and integrates neural inputs - varies hierarchically across the brain. Transmodal cortices, including the DMN, are characterized by longer integration times compared to unimodal regions, underscoring their essential role in higher-order cognitive processin^61^ This hierarchical organization enables the DMN to integrate diverse neural inputs from remote brain regions into its intrinsic signal dynamics, further reinforcing its role as a central hub for complex information processing.^24^ Motivated by our findings that altered temporal dynamics within the DMN can reflect pathological stages, we propose that INT may be related to *in-vivo* EIB. Pathological changes might spread throughout these regions even before cognitive impairment becomes evident.^61^ Indeed, brain regions implicated in the relationship between INT and EIB closely resemble the described topographical connectivity gradient of the DMN,^26^ positioning it as a broader index for brain dynamics influenced by neurotransmission and neurometabolism. Moreover, we show that INT within the DMN is strongly linked to genetic risk factors and cognitive performance, highlighting its potential as a sensitive marker for the early detection of pathological changes.

Consequently, we employed a functional connectome fingerprinting framework to assess the differential identifiability of at-risk individuals. Our results confirm previous reports that AD can be accurately identified based on FC informed by INT,^41^ but also indicate that individuals at genetic risk of AD can be effectively distinguished. Specifically, Idiff is reduced in non-carriers with AD compared to CU carriers, suggesting a narrowing of individual variability among non-carriers as pathology manifests.

Looking more closely at the connections’ distribution, AD-carriers exhibit a greater number of highly identifiable edges, which may reflect a flattening of the brain’s hierarchical organization towards lower area specialization.^41^ In this scenario, brain regions, regardless of their functional specialization, are recruited to compensate for functional decline and maintain overall network operation. This compensatory response, involving excessive whole-brain engagement, may shift a patient’s network organization toward a more random structure, potentially compromising normal information flow.^62^ In contrast, a resilient connectome is thought to selectively engage specific brain regions, with transmodal cortices facilitating long-range interconnections^63,64^ and unimodal cortices remaining largely segregated and specialized.^65^ This dynamic organization helps shape an individual’s unique functional profile.^66^

A spatial reconfiguration of the most identifiable edges was observed in asymptomatic APOE4 carriers, potentially allowing for the identification of at-risk individuals prior to the onset of clinical symptoms. CU APOE4 carriers exhibit an increased emphasis on segregated connections and a reduction in interconnections, regardless of cortical role. This pattern mirrors neurodegenerative processes^65^ characterized by a compensatory attempt at segregation, even in the absence of overt cognitive symptoms. In contrast, AD carriers exhibit widespread modification of identifiable edges with respect to the healthy reference, indicative of a pitting point where neural impairment becomes so severe that brain regions lose their ability to regulate one another effectively.^67,68^ We reveal a picture of the continuum between AD pathology and its genetic risk, where transmodal networks show in both cases a higher number of identifiable connections with respect to controls. However, this does not hold for unimodal cortices showing only specific increase while comparing homogeneous groups (e.g. AD or CU) in the absence of the susceptibility gene. From this point of view, FC informed by INT can be seen as an early surrogate marker of AD in the pathological cascade.^20^ This may further expand our ability to recognize pre-symptomatic and pre-clinical subjects for potential (pharmacological and non-pharmacological) disease-modifying treatments.

However, our findings come with some limitations. Firstly, the *in-vivo* estimation of EIB relies solely on analyzing a unique brain region with single-voxel MRS. Therefore, the inclusion of MRS Imaging (MRSI) in clinical settings would provide further insights allowing for comprehensive whole-brain coverage. Future studies could also consider deriving EIB proxies (i.e. INT) from electrophysiological measures such as EEG and MEG enabling higher temporal richness. The multicentric nature of this study might have intrinsically introduced additional confounding factors, such as variations in MRI acquisition protocols across groups, which however have been carefully evaluated through harmonization.^69^ Additionally, the relatively small sample size and the absence of longitudinal data limit our ability to track changes over time, particularly in predicting which CU subjects carrying the genetic mutation may develop AD and whether the metrics employed could facilitate such predictions. Despite these challenges, this study uniquely enables the multimodal association between *in-vivo* EIB and neurovascular signal derived proxies within a clinical population. This validation supports the use of these proxies as reliable metrics for assessing changes in neural dynamics linked to EIB disruptions. Looking forward, future research should concentrate on understanding the conversion of carriers to elucidate the implications of genetic factors on cognitive impairment and brain function. This could involve larger, longitudinal studies that integrate advanced imaging techniques to capture metabolic, structural, and functional changes over time.

## Conclusion

In conclusion, we non-invasively demonstrated how AD may cause higher-order cortical areas to regress to the functioning of primary cortical areas, resulting in a loss of specialization, both temporally and neuro-metabolically. This regression disrupts the hierarchical continuum of cortical functions, leading to a homogenization that impairs the brain’s ability to process information effectively and downstream impacts cognition. Importantly, these modifications can be observed before cognitive impairment occurs, making them highly valuable for early diagnosis. Indeed, individuals at genetic risk exhibited increased levels of segregation in transmodal networks, potentially indicating an aberrant shifting of FC, which contrasts with the integrative properties typically expected in healthy brain function.

## Data Availability

Data supporting the findings of this study are openly accessible. The analysis code is available on GitHub: https://github.com/ferrariasia06/PATHFIND. For raw material requests related to the Alzheimer's disease cohort (from BrainSync-AD), please contact. Requests concerning raw materials from the cognitively unimpaired subjects cohort (from NEST4AD) should be directed to.

## Acknowledgements

We would like to express our appreciation to Dr. Ryan Raut for the extensive support in data analysis and fruitful discussions about the neuroscientific relevance of these results. We extend our gratitude to Dr. Luca Zigiotto for his invaluable insights into the neuropsychological and cognitive interpretation of the data. His expertise has been instrumental in enhancing our understanding of the findings. We also would like to thank Barbara Cassone for her insights on the functional fingerprinting analysis.

## Supplementary material

Supplementary material is available at *Brain* online.

## Authors contributions

**Francesca Saviola**: Writing - original draft, Formal analysis, Software, Investigation, Writing - review & editing, Conceptualization. **Asia Ferrari**: Formal analysis, Visualization, Software, Writing - review & editing. **Daniele Corbo**: Formal analysis, Visualization, Software, Writing - review & editing. **Michela Pievani**: Formal analysis, Project administration, Writing - review & editing, Funding acquisition. **Silvia Saglia**: Formal analysis, Project administration, Writing - review & editing. **Annamaria Cattaneo**: Resources, Data Curation, Writing - review & editing. **Ilari D’aprile**: Resources, Data Curation, Writing - review & editing. **Giulia Quattrini**: Formal analysis, Project administration, Writing - review & editing. **Valentina Cantoni**: Project administration, Writing - review & editing. **Enrico Premi**: Project administration, Writing - review & editing. **Barbara Borroni**: Writing - review & editing, Project administration, Investigation. **Roberto Gasparotti**: Supervision, Writing - review & editing, Investigation, Supervision, Conceptualization.

---

**Table I.**
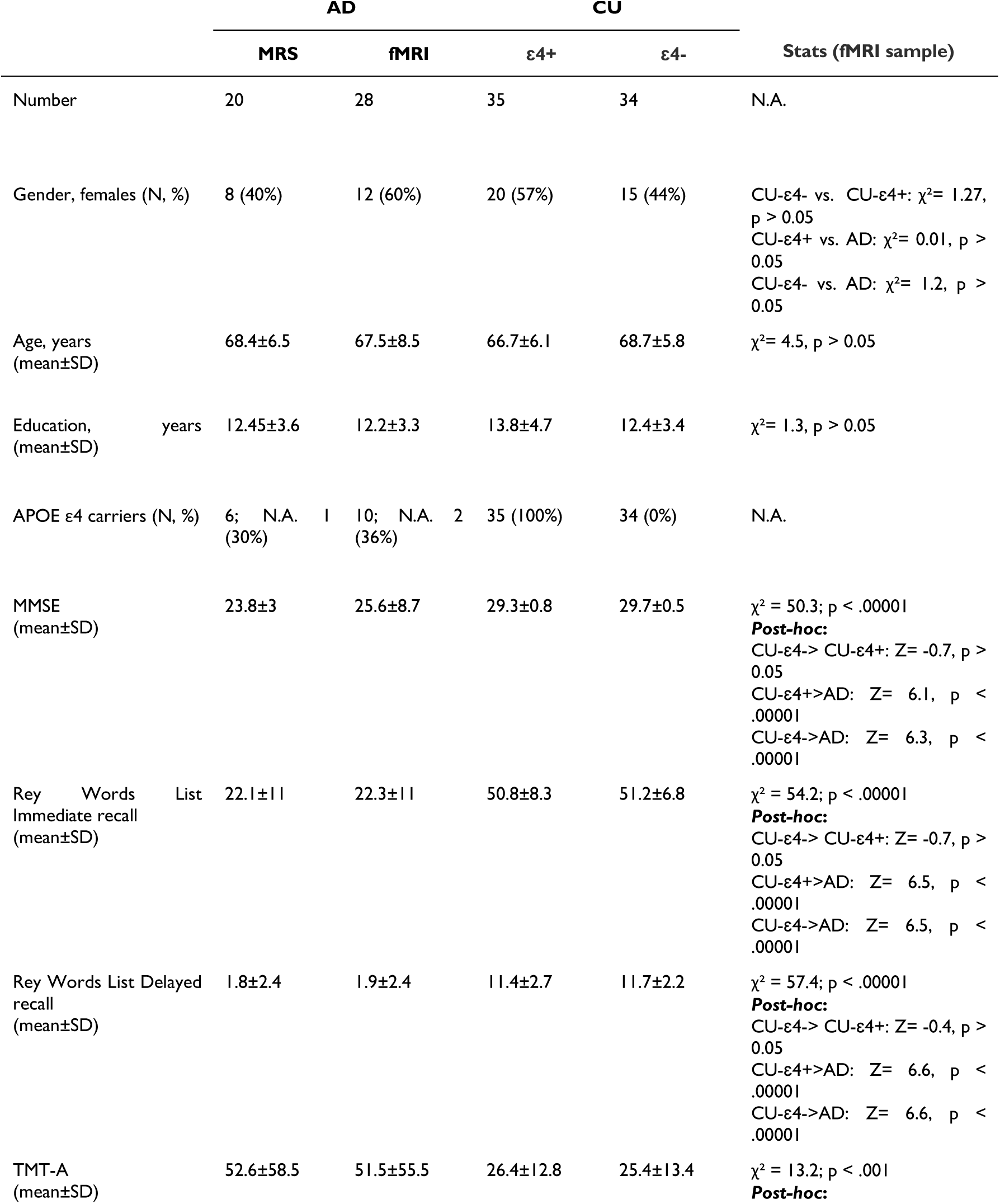
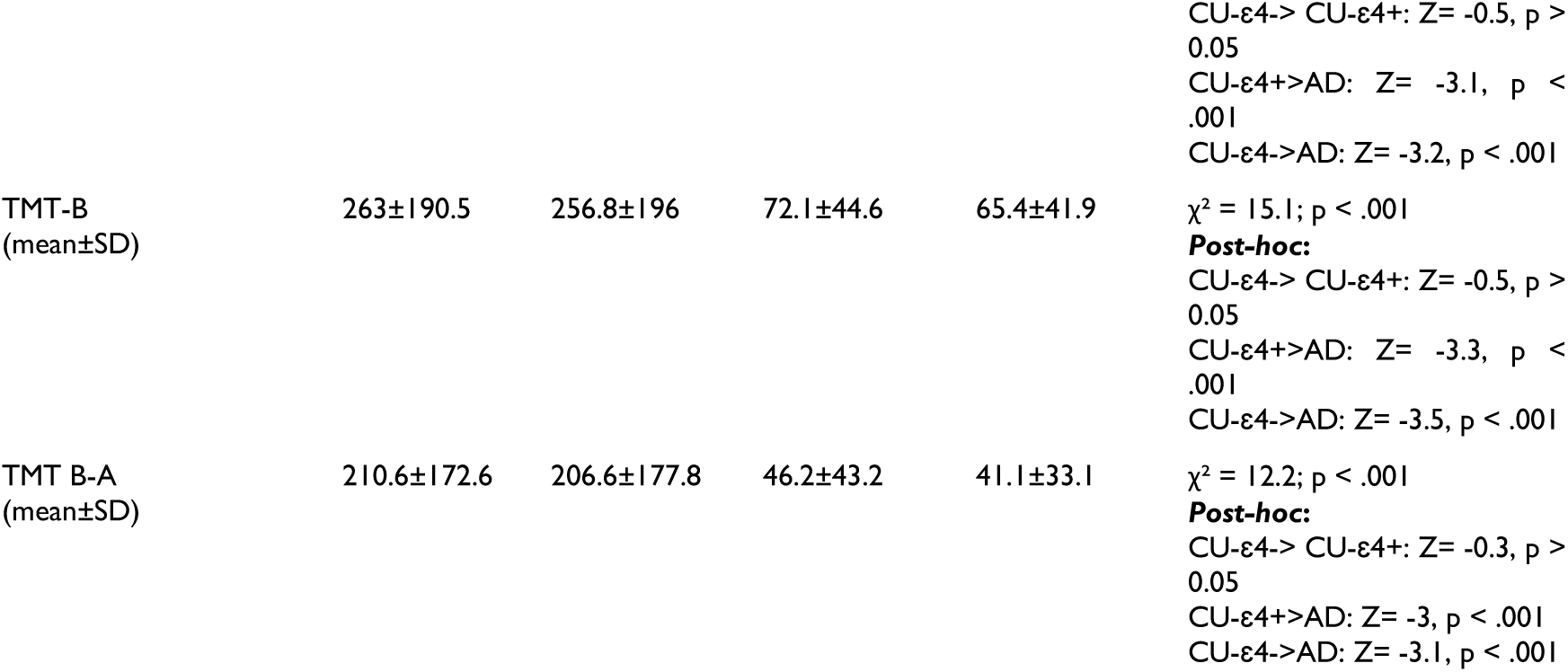
Demographic, genetic, and cognitive characteristics of the samples (AD and CU subjects): Values are reported as mean ± standard deviation (SD) or number (%). χ² and Z denote the test statistics used for the analysis, p the statistical significance (set to p<0.05). Abbreviations: AD (Alzheimer’s disease), CU (Cognitively unimpaired), ɛ4+ (APOE e4 carriers), ɛ4- (APOE e4 non-carriers), MMSE (Mini-Mental State Examination), N.A. (not applicable), TMT-A (Trial Making Test, part A), TMT-B (Trial Making Test, part B), TMT B-A (Trail Making Test, part B minus A), MRS (Magnetic Resonance Spectroscopy), fMRI (functional Magnetic Resonance Imaging).

**Table II.**
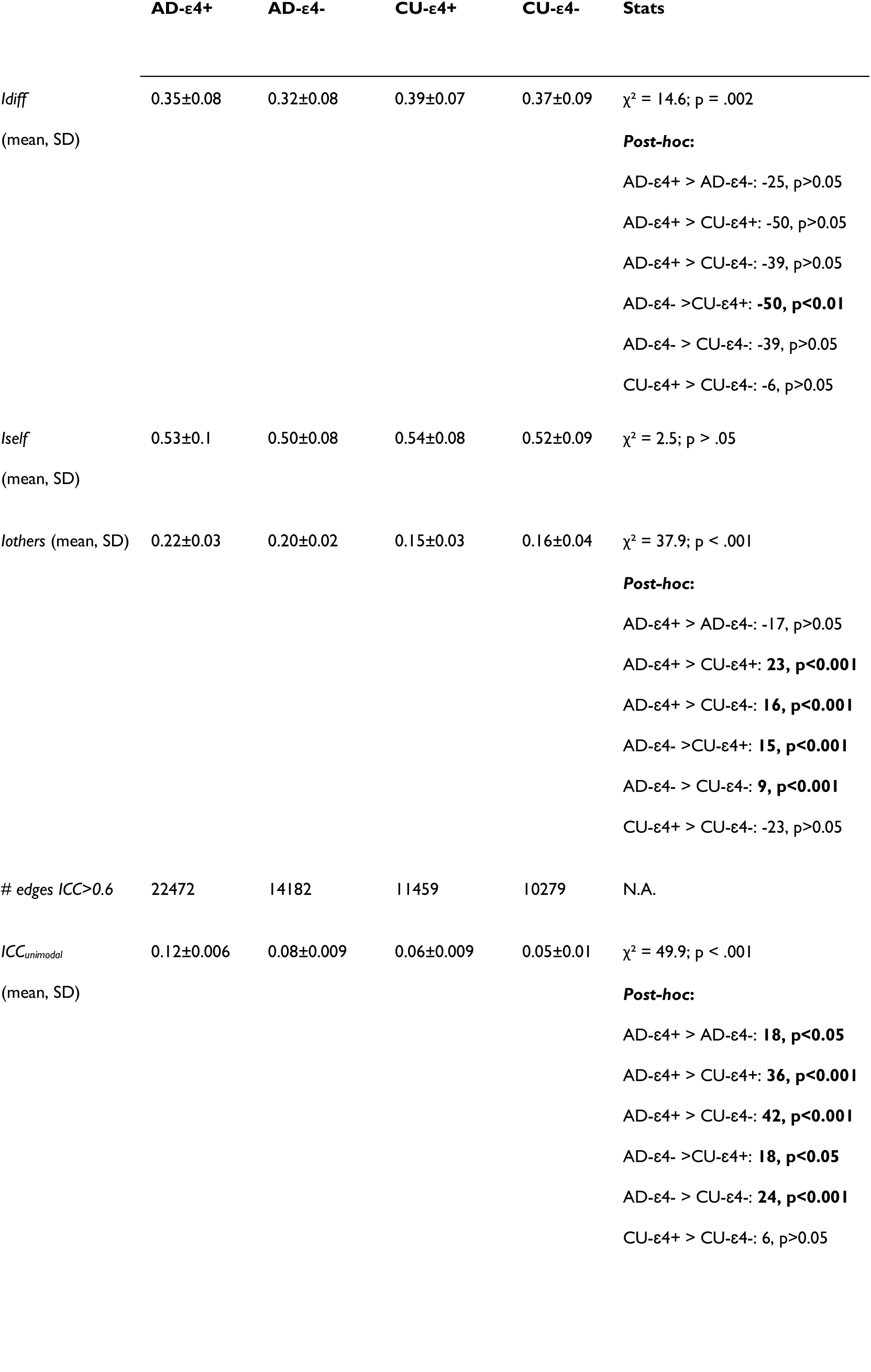
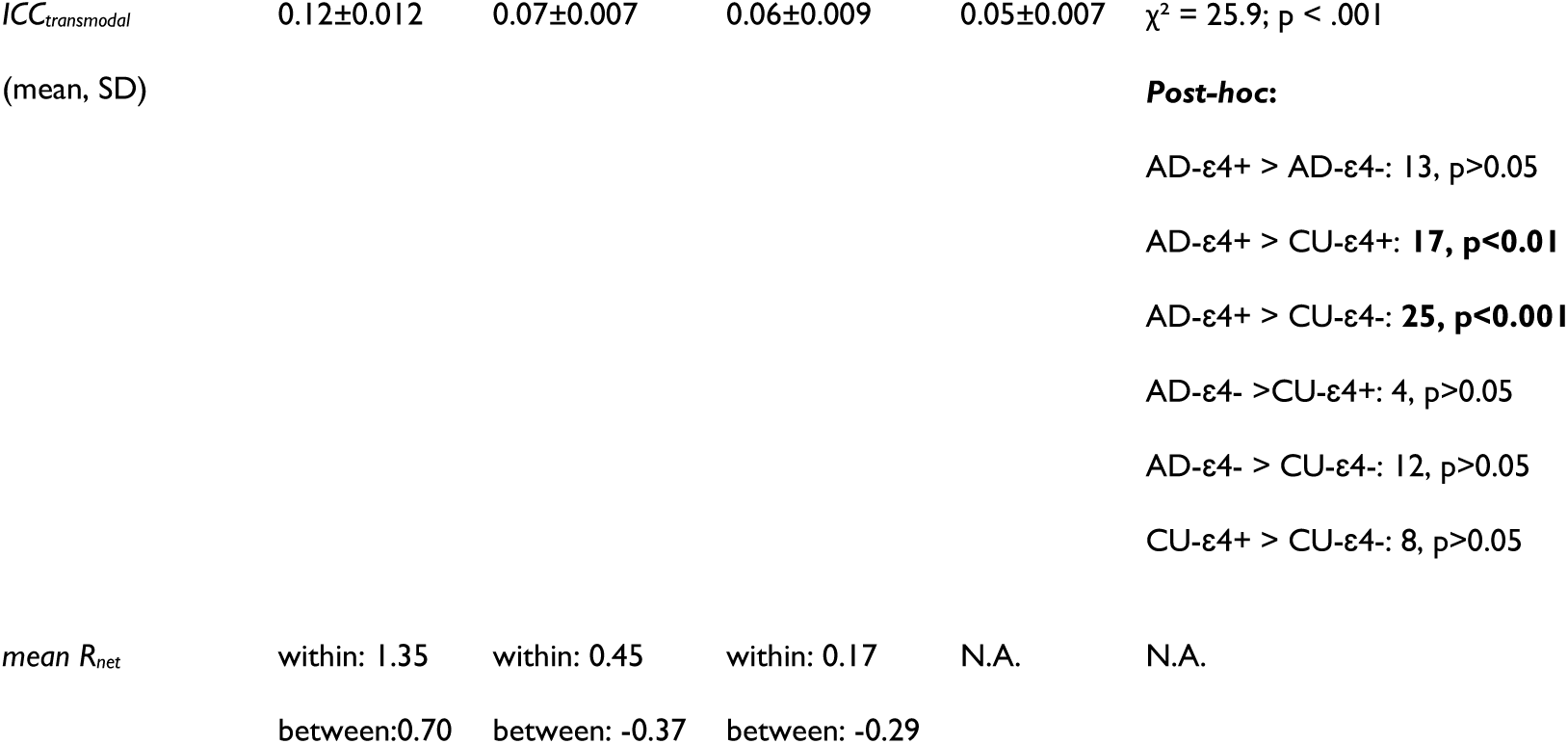
Fingerprinting metrics: For each group, values of the percentage differential identifiability (*Idiff*), self-identifiability (*Iself*), others-identifiability (*Iothers*) are reported. For ICC computation the number of edges with ‘good’ ICC score (i.e. >0.6), mean ‘good’ ICC across network types and mean *Rnet* are reported. Abbreviations: N.A. (Not Applicable).

