## Supplementary Materials for "Unveiling paths to Alzheimer’s disease: excitation-inhibition ratio shapes hierarchical dynamics"

### Methods

#### MR Acquisition protocol

The MR data were acquired with a 3T clinical MR scanner (Skyra, Siemens Healthineers, Erlangen, Germany), equipped with a 64-channel head-neck RF receive coil and parallel transmit RF (operating system VE11C).

For the AD group, the complete protocol was performed in a sample of 20 patients, whereas a shorter version not including MRS acquisition was achieved for further 8 patients (see Table 1). Exclusion criteria specific for MRS encompassed: large nicotine consumption (>1 cigarette/day); large alcohol intake (>14 units/week or >3 days/week) and migraine with aura. Participants were required to abstain from caffeine intake 12 hours pre-scan. Full MR acquisition comprised: (i) T1-weighted MPRAGE (TR=2s, TE=2.85ms, TI=850 ms , 1.1 mm-isotropic voxels); (ii) resting-state fMRI (rs-fMRI) BOLD 2D Echo-Planar Imaging including 200 whole-brain volumes (TR=2.5s, TE=30 ms, 3x3x3.5 mm, full brain coverage, TA=8 min 30 s); (iii) MRS water reference (MEGA-PRESS; TE/TR=68/2000 ms, editing pulses=1.9/7.46 ppm, editing pulses bandwidth=60 Hz, acquisition bandwidth=2000 Hz, cubic voxel size=30 mm3 placed through anatomical landmarks identification in Posterior Cingulate Cortex,[1] spectral points=2048, deactivated VAPOR water suppression, 32 averages, TA=1:20 min);[2–4] (iv) GABA-edited MRS (MEGA-PRESS; TE/TR= 68/2000 ms, brain-optimized automated shimming routine, editing pulses=1.9/7.46 ppm, editing pulses bandwidth=60 Hz, acquisition bandwidth=2000 Hz, same voxel of the MRS water reference, spectral points=2048, VAPOR water suppression, 320 averages, TA=11:06 min).[1]

For APOE ε4+ and ε4- group the acquisition comprised: (i) T1-weighted MPRAGE (TR=2.3s, TE=2 ms, TI=900 ms, 1.0 mm-isotropic voxels); (ii) rs-fMRI BOLD 2D Echo-Planar Imaging and including 600 whole-brain volumes (TR=1s, TE=27 ms, 2.1 mm isotropic voxel, full brain coverage, TA=10 min 15 s).

#### MRS Preprocessing

MRS data were processed with Gannet v3.2.0 (https://github.com/markmikkelsen/Gannet.git) on Matlab version 9.5.0, a Matlab-based analysis tool specifically developed for the analysis of GABA-edited MRS data.[5]

In order to process edited MRS, pre-processing steps included:[6] (i) gross head motion correction; and (ii) phase and frequency drift correction with a robust spectral registration algorithm.[7] After these corrections, the Fourier transform is computed to perform the frequency spectra subtraction between the ON and OFF spectra, which are aligned and averaged.[5] Once the pre-processing is performed, the neuro-metabolite spectra peaks are extracted by peak integration fitting targeting GABA+ and Glx (a compound metabolite of Glutamate+Glutamine) in the difference spectrum.

The fitted spectra were then corrected by considering the proportion of different tissue types in the voxel, namely gray matter (GM), white matter (WM), and CSF. To do this, the single voxel was co-registered on the subject T1-weighted anatomical image with SPM12 (Statistical Parametric Mapping, n.d. https://www.fil.ion.ucl.ac.uk/spm/software/spm12/) and then segmented in different tissue types. Concentrations of metabolites of interest were quantified relative to the internal water reference.[5] Therefore, the unitless signal intensity values are converted into absolute concentrations with a computation weighting for the percentage of the tissue type involved[8] enabling comparison between different subjects.

Finally, the macromolecules (MM) contribution was considered by using the Gannet model,[6] which assumes that 45% of the area of GABA+ peak is attributable to MM contribution.[5] In order to evaluate neuro-metabolite concentration changes in terms of EIB balance. The overall average neuro-metabolite concentration was averaged across each full MRS acquisition session in Gannet (GABAGlx model) and reported as units of concentration.

#### MRS quality control

Data exclusion criteria were established for the processing methods, taking into account the type of fitting procedure. Quality metrics include: GABA Signal-to-noise ratio (SNR) and fit error, Glx SNR and fit error.[6,9] Two subjects were excluded due to errors in the voxel placement during acquisition and high extent of head motion (Fig. 2) whereas one subject was excluded due to poor spectral quality.

### MRI Preprocessing and quality assurance

A standard preprocessing pipeline will be performed using the FSL software.[10] The preprocessing includes: (1) slice timing correction; (2) T1-weighted image tissue segmentation; (3) geometric distortion and head motion correction with DVARS deweighting;[11,12] (4) co-registration of the T1-weighted image to the time-series, (5) nuisance regressions of the mean white matter signal, the mean cerebrospinal fluid signal and the six head motion parameters; (6) band-pass temporal filtering [0.01-0.1 Hz]; (7) normalization to standard MNI template space; and (8) spatial smoothing 6 mm FWHM Gaussian kernel size. To ensure the quality of pre-processed fMRI data, we established criteria for excluding volumes based on Framewise Displacement (FD) and DVARS. We set a threshold of 0.5 mm for FD[13,14] to exclude volumes with excessive motion.[11] For DVARS, we applied the FSL standard method, which calculates the temporal derivative of root mean square (RMS) intensity changes to assess frame-to-frame variations. Using a run-specific approach, we determined a cutoff based on the distribution of DVARS values across sessions, excluding frames that exceeded 5%. Additionally, the mean tSNR value was compared between groups to assess the signal strength.

### CU-ε4+/CU-ε4- recruitment

The cognitively unimpaired (CU) group recruitment involved a comprehensive neuropsychological evaluation. Participants were included in the study only if they did not demonstrate deficits in two or more cognitive tests. This evaluation incorporated assessments of global cognition using the Mini-Mental State Examination (MMSE); Folstein et al., 1975). Memory was evaluated through the Rey Auditory Verbal Learning Test, assessing both immediate and delayed recall,[15,16] as well as Story Recall[17] and the Rey–Osterrieth Complex Figure recall.[18–20] To assess visuospatial abilities, participants completed the Rey-Osterrieth Complex Figure copy task.[20] Attention and executive functions were measured using the Digit Span Forward Test,[21] the Trail Making Test parts A and B,[22–24] and the Symbol Digit Test.[23] Language abilities were evaluated through phonemic and semantic verbal fluency tests.[17]

### Quality assurance

#### Head motion

Given the critical role of head shifts in analyzing INT as highlighted by Goldberg et al.,[25] our methodology incorporated several preprocessing steps to address geometric distortions and correct for head motion. Initially, we employed DVARS deweighting[11] to mitigate the impact of motion artifacts on the data quality. To quantitatively assess head motion throughout each run, we calculated the framewise displacement (FD).[11] This FD was subsequently integrated into our linear models to evaluate its potential influence on the observed effects. By examining whether FD could account for variability in INT estimates, we aimed to discern if head motion significantly affected our findings.

**Table 1: Data Quality Metrics for fMRI Data. This table presents the data quality metrics for the final sample included in the analysis, comprising 28 Alzheimer's disease (AD) participants, 35 ε4- individuals, and 34 ε4+ individuals. Abbreviations: FD (Framewise displacement), DVARS (Derivative of Variance over voxels), tSNR (temporal signal to noise ratio)**

| | **Group (mean±SD; [range])** | | | **Statistical testing (p-value, F-value)** |
|---|---|---|---|---|
| | **ε4-** | **ε4+** | **AD** | |
| Translation (mm) | 0.0002±0.002; [0 0.06] | 0.0002±0.001; [0 0.03] | 0.00002±0.001; [0 0.06] | >0.05 |
| Rotation (gradient) | -0.001±0.07; [0 2.1] | -0.004±0.08; [0 3] | -0.01±0.1; [0 2.8] | >0.05 |
| FD | 0.05±0.04; [0 0.5] | 0.04±0.03; [0 0.4] | 0.07±0.06; [0 1] | >0.05 |
| DVARS | 59.7±10.8; [0 234.2] | 55.6±9.3; [0 179.3] | 41±18.6; [0 314.1] | **<0.05; F-value=6.4** |
| tSNR | 31.70±5.24 | 32.07±6.49 | 58.84±8.44 | **<0.01; F-value=5.0** |

#### Temporal SNR

The analysis of tSNR values across different functional networks revealed notable variations between groups, as indicated by the Kruskal-Wallis tests, which resulted in a $p < 0.01$ for all networks examined. In the Visual network, the post-hoc Wilcoxon rank-sum test for the comparison between AD and ε4+ indicated a highly significant result ($p < 0.01$, $z = 5.28$), while the comparison between AD and ε4- also demonstrated strong evidence of difference ($p < 0.01$, $z = 5.68$). However, no meaningful difference was found between ε4+ and ε4- ($p = 0.1230$, $z = 1.54$). In the Somatomotor network, there were clear distinctions for the comparison between AD and ε4+ ($p < 0.01$, $z = 5.61$) as well as for AD versus ε4- ($p < 0.01$, $z = 5.99$). The comparison between ε4+ and ε4- did not reveal significant results ($p = 0.1808$, $z = 1.34$). The Dorsal Attention network also showed marked differences, with AD compared to ε4+ yielding ($p < 0.01$, $z = 4.20$) and AD compared to ε4- showing ($p < 0.01$, $z = 4.87$). The comparison between ε4+ and ε4- approached significance but did not reach it ($p = 0.0747$, $z = 1.78$). In the Ventral Attention network, substantial differences were observed for AD versus ε4+ ($p < 0.01$, $z = 6.56$) and AD versus ε4- ($p < 0.01$, $z = 6.64$), while no notable difference was found between ε4+ and ε4- ($p = 0.4604$, $z = 0.74$). The Limbic network results reflected similar trends with clear distinctions between AD and ε4+ ($p < 0.01$, $z = 4.75$) and AD and ε4- ($p < 0.01$, z = 5.37), but no significant difference was noted between ε4+ and ε4- ($p = 0.1620$, $z = 1.40$). In the Frontoparietal network, comparisons yielded strong evidence for differences between AD and ε4+ ($p < 0.01$, $z = 5.40$) as well as for AD versus ε4- ($p < 0.01$, $z = 5.79$). The comparison between ε4+ and ε4- showed no statistically meaningful results ($p = 0.5130$, $z = 0.65$). Finally, in the Default Mode network, significant findings were reported for both comparisons: AD versus ε4+ ($p < 0.01$, $z = 6.20$) and AD versus ε4- ($p < 0.01$, $z = 6.32$), while no substantial difference was observed between ε4+ and ε4- ($p = 0.3105$, $z = 1.01$). These results highlight the distinct tSNR profiles across different networks concerning the varying groups studied.

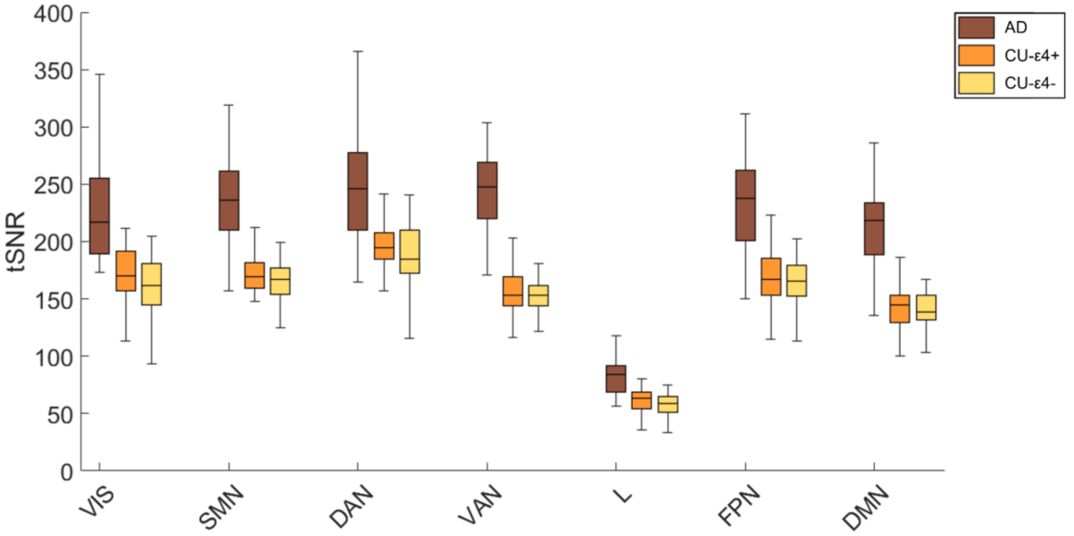


**Figure 1: Temporal Signal-to-Noise Ratio (tSNR) Values Across Yeo Networks for the three groups.** Abbreviations: VIS: Visual, SMN: Somatosensory, DAN: Dorsal Attention, VAN: Ventral Attention, L: Limbic, FPN: Fronto-Parietal, DMN: Default Mode Network.

**Table 2: Temporal Signal-to-Noise Ratio (tSNR) values across functional networks. This table presents the mean ± standard deviation of tSNR values for the different groups (AD, CU-ε4+, CU-ε4−) groups across Yeo functional networks. These metrics provide a group-wise overview of tSNR values, highlighting network-specific differences. Abbreviations: VIS: Visual, SMN: Somatosensory, DAN: Dorsal Attention, VAN: Ventral Attention, L: Limbic, FPN: Fronto-Parietal, pDMN: Posterior Default Mode Network, aDMN: Anterior Default Mode Network.**

| | **AD** | **CU-ε4+** | **CU- ε4-** |
|---|---|---|---|
| VIS | 223.84 ± 40.47 | 171.31 ± 22.66 | 160.12 ± 28.11 |
| SMN | 234.66 ± 39.58 | 172.98 ± 20.49 | 163.80 ± 21.18 |
| DAN | 251.67 ± 51.73 | 200.33 ± 30.28 | 185.81 ± 30.38 |
| VAN | 243.49 ± 34.25 | 156.81 ± 19.73 | 150.48 ± 21.19 |
| L | 82.89 ± 17.09 | 60.02 ± 12.89 | 56.46 ± 12.58 |
| FPN | 234.76 ± 41.27 | 169.20 ± 28.49 | 161.46 ± 26.35 |
| DMN | 213.19 ± 35.46 | 142.72 ± 20.72 | 136.48 ± 21.77 |

#### Brain behavior relationship

PLS-C is a multivariate statistical method that finds the latent variables, or mutually orthogonal, weighted linear combinations of the original variables in the two datasets that have the highest degree of correlation with one another. In the current analysis, one dataset represents the INT values of the DMN (i.e., $X_{n\times t}$) with n=97 rows as the sample size of all subjects, and t=101 columns as the number of parcels of INT within the DMN. The other dataset encompasses cognitive performance (i.e. attention and executive scores: Trail Making Test (TMT-A, TMT-B, TMT B-A); memory scores: immediate and delayed Rey word's list) representing behavioral variables (i.e., $y_{n\times m}$) with n=97 rows as the sample size and m=11 columns as the number of behavioral variables (i.e. (1) presence of ε4 (0/1), (2-4) TMT score, (5) 15-words Rey's list immediate and (6) delayed word recall, (7-11) with relative interactions with ε4). Whitin data matrices and behavioral variables were normalized column-wise (i.e., z-scored across the population) while interactions were computed as multiplication between z-scored variables to identify the latent variables. The correlation matrix R=X'Y was then subjected to the following singular value decomposition: R=X'Y=USV' where $S_{m\times m}$ is the diagonal matrix of singular values and $U_{t\times m}$ and $V_{m\times m}$ are the orthonormal matrices of the left and right singular vectors, respectively. A latent variable corresponds to each column in the **U** and **V** matrices. Each element of the diagonal of **S** is the corresponding singular value. The DMN INT features' and behavioral features' relative contributions to latent variables are shown by the left and right singular vectors, U and V, respectively.

Positively weighted DMN INT features correlate with positively weighted behavioral features, whereas negatively weighted DMN INT and behavioral correlate with each other. Brain scores show how much each area of the brain within the DMN displays the weighted patterns found by latent variables that can be estimated using singular vectors. The computation of brain scores for INT in the DMN and behavioral characteristics involves projecting the initial data onto the weights that are determined from PLS, specifically **U** and **V**, obtaining:

- Brain scores for DMN INT features = **XU**
- Brain scores for behavioral features= **YV**

The Pearson correlation coefficient between the original data matrices and the relevant brain scores are then used to calculate loadings for DMN INT features and behavioral features. The correlation coefficients between the initial DMN INT characteristics vectors and the PLS-derived brain scores for DMN INT features, for instance, are known as temporal DMN INT features’ loadings. Using 10000 permutation tests, the statistical significance of latent variables (LC) was evaluated. The original data was randomized using spatial autocorrelation-preserving nulls. Every permutation was subjected to the PLS analysis once more, producing a null distribution of singular values. After that, the original singular values' significance was evaluated in comparison to the permuted null distributions. Using bootstrap resampling, which involves randomly resampling rows of the original data matrices **X** and **Y** 500 times with replacement, the dependability of PLS loadings was assessed. Next, for every resampled data set, the PLS analysis was performed once again to provide a sampling distribution for every DMN INT feature and behavioral feature (i.e., 500 bootstrap-resampled loadings). We next utilize the bootstrap-resampled loading distributions to determine the loadings' 95% confidence intervals (Fig. 3C). The decomposition of each LC in the set of behavioral weights and DMN INT features weights, represents how largely each variable contributes to the multivariate brain-behavior correlations across runs.

#### Linear Models

We conducted tests to identify the best-fitting statistical model for the INT data. Initially, we evaluated several model types, including inverse Gaussian, Gamma, and Weibull distributions. Model performance was assessed using the Akaike Information Criterion (AIC) and Bayesian Information Criterion (BIC). Ultimately, the log-normal model was determined to be the most appropriate, as it exhibited lower AIC and BIC values compared to the other models. Additionally, applying a log transformation to the dependent variable effectively addressed data skewness, improving both the interpretability and robustness of the results.

### Results

#### Clinical scores

**Table 3: Clinical Scores for the Alzheimer's Disease group. This table summarizes the clinical scores for the Alzheimer's disease (AD) group, for both the edited-MRS and the fMRI subsamples. The metrics include mean values (M) and standard deviations (SD) for two assessments: the Clinical Dementia Rating (CDR) sum of boxes and the Alzheimer’s Disease Assessment Scale-Cognitive Subscale (ADAS-Cog13).**

| | **edited-MRS** | **fMRI** | **Stats** |
|---|---|---|---|
| Number | 19 | 28 | N.A. |
| Gender | 7 females | 12 females | $\chi^2$ = 0.2, $p$ > 0.05 |
| Age (mean±SD) | 68.4±6.5yo | 67.5±8.5yo | $\chi^2$ = 0.1, $p$ > 0.05 |
| Education (mean±SD) | 12.45±3.6y | 12.2±3.3 | $\chi^2$ = -0.5, $p$ > 0.05 |
| MMSE (mean±SD) | 23.8±3 | 25.6±8.7 | $\chi^2$ = -0.2, $p$ > 0.05 |
| Rey Words Immediate (mean±SD) | 22.1±11 | 22.3±11 | $\chi^2$ = -0.3, $p$ > 0.05 |
| Rey Words Delayed (mean±SD) | 1.8±2.4 | 1.9±2.4 | $\chi^2$ = -0.8, $p$ > 0.05 |
| TMT-A (mean±SD) | 52.6±58.5 | 51.5±55.5 | $\chi^2$ = 0.2, $p$ > 0.05 |

| | | | |
|---|---|---|---|
| TMT-B (mean±SD) | 263±190.5 | 256.8±196 | $\chi^2 = 0.4$, $p > 0.05$ |
| TMT B-A (mean±SD) | 210.6±172.6 | 206.6±177.8 | $\chi^2 = 0.6$, $p > 0.05$ |
| CDR sum of boxes (mean±SD) | 2.4±1.5 | 2.4±1.5 | $\chi^2 = 0.1$, $p > 0.05$ |
| ADASCog13 (mean±SD) | 17.83±5.5 | 17.87±4.9 | $\chi^2 = -0.3$, $p > 0.05$ |

### MRS

**Table 4: Data quality of GABA-edited MRS fitted in Gannet. The table shows data quality in the final utilized sample ($N_{QA}$=19).**

| QA properties | Participants ($N_{QA}$=19) |
|---|---|
| GABA SNR (mean±SD) | 21.63±5.36 |
| GABA Fit error (mean±SD) | 4.27±1.10 |
| GABA FWHM (mean±SD) | 19.86±1.45 |
| Glx SNR (mean±SD) | 34.69±7.59 |
| Glx Fit error (mean±SD) | 2.67±0.76 |
| Glx FWHM (mean±SD) | 14.22±0.71 |

#### MRS Voxel placement

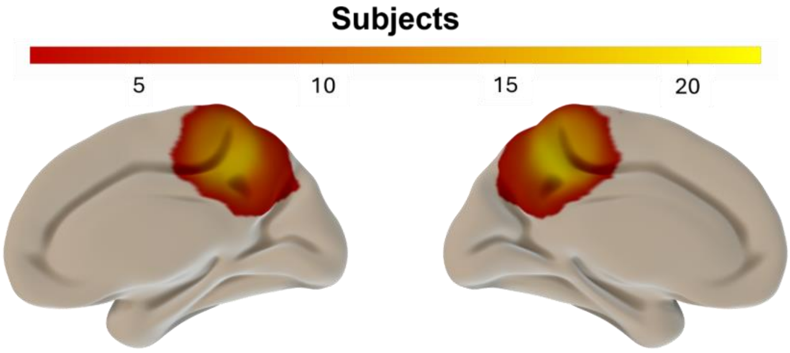


**Figure 2: Overall voxel placement across subjects.**

#### Association of EIB with clinical scores

Further results from the GAMLSS, which analyzed clinical score as the dependent variable and included a fixed-effect factor for neurometabolites, highlighted a non-significant trend of GABA in association with ADAS-Cog ($\beta$ = -0.29, $p_{FDR}$ = 0.068). These findings indicate that while these results may not have reached statistical significance, the significant effect observed for EIB could be driven by GABA.

#### INT as a proxy measure for E/I balance

In the Alzheimer's disease cohort, INT patterns exhibited a voxel-wise positive relationship with compound neurometabolites GABA+ and Glx. The spatial distribution of significant INT cluster peaks associated with the neurometabolites was predominantly localized within the Frontal Medial Cortex (GABA+: $t$ = 13.61; voxel-wise TFCE $p_{FWE} < 0.02$; Glx: $t$ = 13.67; voxel-wise TFCE $p_{FWE} < 0.02$), which belong to the anterior portion of the DMN. Similar results were observed for the, as detailed in the supplementary materials.

**Table 5: Whole-brain INT patterns associated with EIB, GABA+ and Glx. This table summarizes the whole-brain intrinsic neural timescale (INT) patterns associated with the neuro-metabolites GABA+ and Glx, and their ratio. For each metabolite, the table reports the significant peak, detailing anatomical areas as defined by the Harvard-Oxford Cortical Structural Atlas, MNI coordinates, T-values, and family-wise error (FWE) corrected p-values (PFWE).**

| Neurometabolite / ratio | Area | MNI coordinates | | | T-val | $P_{FWE}$ |
|---|---|---|---|---|---|---|
| | | x | y | z | | |
| | Frontal Medial Cortex | -6 | 38 | -20 | 11.89 | 0.002 |
| | Frontal Medial Cortex; Paracingulate Gyrus | -12 | 38 | -12 | 11.89 | 0.002 |
| | Frontal Orbital Cortex; Temporal Pole | -26 | 18 | -26 | 11.28 | 0.002 |
| | Frontal Orbital Cortex | -18 | 22 | -26 | 11.28 | 0.002 |
| EIB | Frontal Orbital Cortex; Frontal Pole | -16 | 30 | -26 | 11.28 | 0.002 |
| | Frontal Orbital Cortex | -18 | 18 | -24 | 11.28 | 0.002 |
| | Frontal Medial Cortex; Paracingulate Gyrus | -12 | 38 | -12 | 13.61 | 0.002 |
| | Frontal Medial Cortex | -6 | 38 | -20 | 13.61 | 0.002 |
| | Frontal Orbital Cortex; Insular Cortex | 32 | 22 | -12 | 12.33 | 0.002 |
| | Frontal Orbital Cortex; Insular Cortex | 28 | 22 | -12 | 12.33 | 0.002 |
| GABA+ | Frontal Pole | -6 | 70 | -2 | 12.33 | 0.002 |
| | Frontal Pole | -12 | 50 | -26 | 12.33 | 0.002 |
| | Frontal Medial Cortex | -6 | 38 | -20 | 13.67 | 0.002 |
| | Frontal Medial Cortex; Paracingulate Gyrus | -12 | 38 | -12 | 13.67 | 0.002 |
| | Frontal Orbital Cortex; Temporal Pole | -26 | 18 | -26 | 13.55 | 0.002 |
| | Frontal Orbital Cortex | -18 | 22 | -26 | 13.55 | 0.002 |
| Glx | Frontal Orbital COrtex; Frontal Pole | -16 | 30 | -26 | 13.55 | 0.002 |
| | Frontal Orbital COrtex | -18 | 18 | -24 | 13.55 | 0.002 |

### ACF decay in the Default Mode Network

We replicated the ACF decay analysis first on the whole DMN, and subsequently on the anterior and posterior DMN. The Kruskal-Wallis test revealed significant differences in ACF decay among groups for the whole DMN ($\chi^2$ =8.2, $p$ =0.02), with post-hoc comparisons showing that the AD group significantly differed from both ε4+ (*Cohen's d* =-0.05, $z$ =-2.3, $p$ = 0.02) and ε4- (*Cohen's d* =-0.07, $z$ = -2.83, $p$ = 0.005), while no difference was found between ε4+ and ε4- ($z$ = -0.79, $p$ > 0.05). Similar results were observed in the anterior ($\chi^2$ = 6.7, $p$ =

0.03) and posterior ($\chi^2$ = 9.9, *p* = 0.007) DMN regions, where Wilcoxon rank-sum tests confirmed significant differences for the AD group compared to both ε4+ and ε4- groups in each region. Specifically, in the anterior DMN, AD vs. ε4+: *Cohen's d* =-0.05, *z* =-2, *p* = 0.048; AD vs. ε4-: *Cohen's d* = -0.06, *z* = -2.6, *p* = 0.001; and in the posterior DMN, AD vs. ε4+: *Cohen's d* = -0.07, *z* = -2.7, *p* = 0.007; AD vs. ε4-: *Cohen's d* = -0.07, *z* = -3.1, *p* = 0.002. No significant differences were observed between ε4+ and ε4- groups in either DMN region. Thus, DMN ACF decay is significantly altered in AD patients compared to both carriers and non-carriers, with no difference between the ε4 groups.

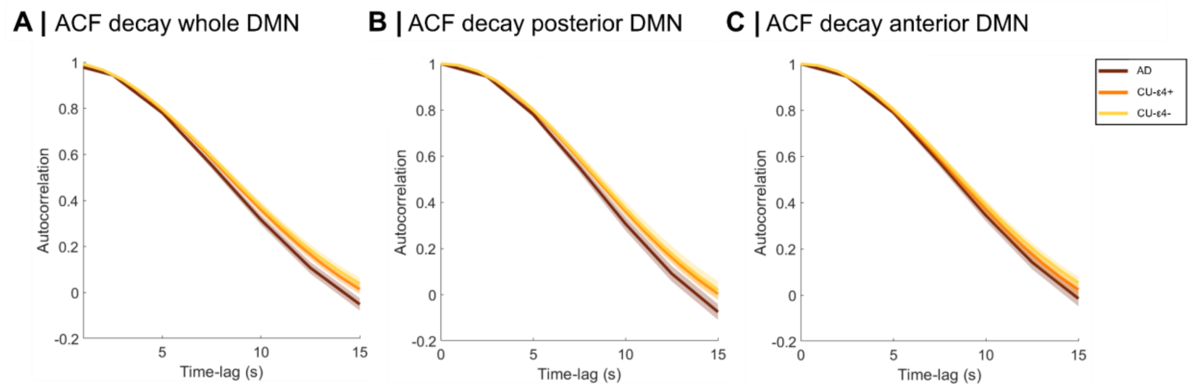


**Figure 3: DMN ACF decay across groups.** (**A**) ACF decay of the Default Mode network (DMN). (**B**) ACF decay of the posterior portion of the DMN**.** (**C**) ACF decay of the anterior portion of the DMN.

#### Intrinsic Neural Timescal group differences

INT across groups revealed significant differences in the DMN and the hippocampal parcels (whole DMN: $\chi^2$ = 130.9, *p* < 0.0001, anterior DMN: $\chi^2$ = 68.5, *p* < 0.0001, posterior DMN: $\chi^2$ = 122.3, *p* < 0.0001, hippocampus: $\chi^2$ = 28.9, *p* < 0.0001). Significant pairwise differences between AD and ε4+ groups, AD and ε4- groups, and ε4+ and ε4- groups are found across all regions (*p* < 0.05). A further stratification based on cognitive impairment (AD/CU) and gene mutation status (ε4+/ε4-) allowed us to test differences in INT across the four groups. Pairwise comparisons using Wilcoxon rank-sum tests indicated significant differences between AD+ and AD- groups (whole DMN: *d* = 0.06, *z* = 3, *p* = 0.003; posterior DMN: *d* = 0.07, *z* = 2.3, *p* = 0.02; hippocampus: *d* = 0.1, *z* = 2.2, *p* = 0.03), AD+ and CU+ groups (whole DMN: *d* = -0.08, *z* = -5.5, *p* < 0.0001; anterior DMN: *d* = -0.06, *z* = -2.8, *p* = 0.005; posterior DMN: *d* = -0.1, *z* = -5.1, *p* < 0.0001), AD+ and CU- groups (whole DMN: *d* = -0.1, *z* = -8, *p* < 0.0001; anterior DMN: *d* = -0.1, *z* = -5.1, *p* < 0.0001; posterior DMN: *d* = -0.1, *z* = -6.3, *p* < 0.0001;

hippocampus: $d = -0.1$, $z = -2.5$, $p = 0.01$), AD- and CU+ groups (whole DMN: $d = -0.2$, $z = -10.8$, $p < 0.0001$; anterior DMN: $d = -0.1$, $z = -6.2$, $p < 0.0001$; posterior DMN: $d = -0.2$, $z = -9.4$, $p < 0.0001$; hippocampus: $d = -0.2$, $z = -4.7$, $p < 0.001$), AD- and CU- groups (whole DMN: $d = -0.2$, $z = -13.8$, $p < 0.0001$; anterior DMN: $d = -0.2$, $z = -9$, $p < 0.0001$; posterior DMN: $d = -0.2$, $z = -10.7$, $p < 0.0001$; hippocampus: $d = -0.2$, $z = -6.2$, $p < 0.001$), CU+ and CU- groups (whole DMN: $d = -0.1$, $z = -4.2$, $p < 0.0001$; anterior DMN: $d = -0.1$, $z = -3.6$, $p < 0.001$; posterior DMN: $d = -0.04$, $z = -2.2$, $p = 0.03$).

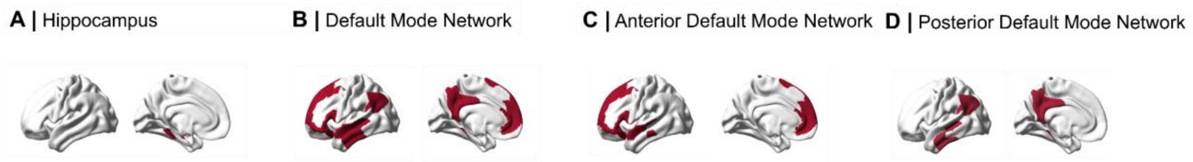


**Figure 4: Regions of interest.** (**A**) Hippocampus. (**B**) Default Mode network (DMN). (**C**) Anterior portion of the DMN**.** (**D**) Posterior portion of the DMN.

### Linear Models

In our analysis investigating the relationship between *Sex* and INT, while controlling for framewise displacement,[11] we identified a significant fixed effect in the posterior DMN ($\beta = 0.033$, $p_{FDR} = 0.02$), and a non-significant trend for the whole DMN ($\beta = 0.023$, $p_{FDR} = 0.05$).
The analysis assessing linear relationships between INT and the independent variables *Group* and *Sex*, performed on the hippocampus revealed a fixed effect of the variable *Group* ($\beta = -0.09$, $p_{FDR} < 0.01$). Whereas the linear models investigating the link between hippocampal INT values and MMSE, showed a significant fixed effect ($\beta = 0.008$, $p_{FDR} = 0.03$).
The analysis revealed that the linear models incorporating both *Sex* and MMSE as independent variables had higher Bayesian Information Criterion (BIC) and Akaike Information Criterion (AIC) values compared to the model that included only the MMSE. This indicates that the model with only the MMSE better explains the data.
Furthermore, all models reported in the main manuscript were re-evaluated with framewise displacement included as a covariate, acknowledging its potential confounding influence within the Intrinsic Neural Timescale framework.[25]. However, this confounding variable did not yield significant results in any of the models. The inclusion of this method was solely to control for motion-related confounds, as deweighting of DVARS was performed during preprocessing steps. More in detail, the models investigating the relationship between INT and the independent variables *Sex* and *Group* did not show significant effects for the framewise

displacement: DMN ($\beta$ = -0.3, $p_{FDR}$ = 0.2), posterior DMN ($\beta$ = -0.4, $p_{FDR}$ = 0.1), anterior DMN ($\beta$ = -0.2, $p_{FDR}$ = 0.4), hippocampus ($\beta$ = 0.03, $p_{FDR}$ = 0.9). Moreover, the models investigating the relationship between INT and the MMSE also did not show significant effects for the framewise displacement: DMN ($\beta$ = -0.3, $p_{FDR}$ = 0.3), posterior DMN ($\beta$ = -0.4, $p_{FDR}$ = 0.4), anterior DMN ($\beta$ = -0.2, $p_{FDR}$ = 0.5), hippocampus ($\beta$ = 0.02, $p_{FDR}$ = 0.9).

### Differences across groups correspondence to Yeo functional network

**Table 6: Whole-brain INT differences across AD, ε4+, and ε4-. This table summarizes whole-brain intrinsic neural timescale (INT) patterns that differ between groups divided for functional networks as defined by the Yeo Atlas. Abbreviations: VIS: Visual, SMN: Somatosensory, DAN: Dorsal attention, VAN: Ventral attention, L: Limbic, FPN: Fronto-parietal, pDMN: posterior Default mode network, aDMN: anterior Default mode network.**

| Contrast | VIS | SMN | DAN | VAN | L | FPN | pDMN | aDMN |
|---|---|---|---|---|---|---|---|---|
| ε4- > AD | 17.7% | 13.6% | 9.3% | 12.8% | 8.7% | 10.7% | 12.2% | 15.1% |
| ε4+ > AD | 18.8% | 13% | 9.6% | 11.4% | 8.3% | 11.4% | 12.9% | 14.5% |
| ε4- > ε4+ | 15.6% | 13.4% | 9.7% | 13.4% | 10.4% | 10.4% | 11.2% | 16% |

### Permutation testing fingerprinting

The results of the Wilcoxon rank sum tests comparing Idiff values against their corresponding null distributions revealed significant differences across all groups (AD-ε4+: $z$ = -5.4446, $p$ < 0.01; AD-ε4-: $z$ = -6.8696, $p$ < 0.01; ε4+: $z$ = -10.0671, $p$ < 0.01; ε4-: $z$ = -9.927, $p$ < 0.01). When examining SR, we observed similarly significant results (AD-ε4+: $z$ = -5.7643, $p$ < 0.01; AD-ε4-: $z$ = -7.2612, $p$ < 0.01; ε4+: $z$ = -10.5369, $p$ < 0.01; ε4-: $z$ = -10.4463, $p$ < 0.01). These findings indicate that both Idiff and SR values for all groups significantly diverge from their respective null distributions.

### ICC statistical analysis

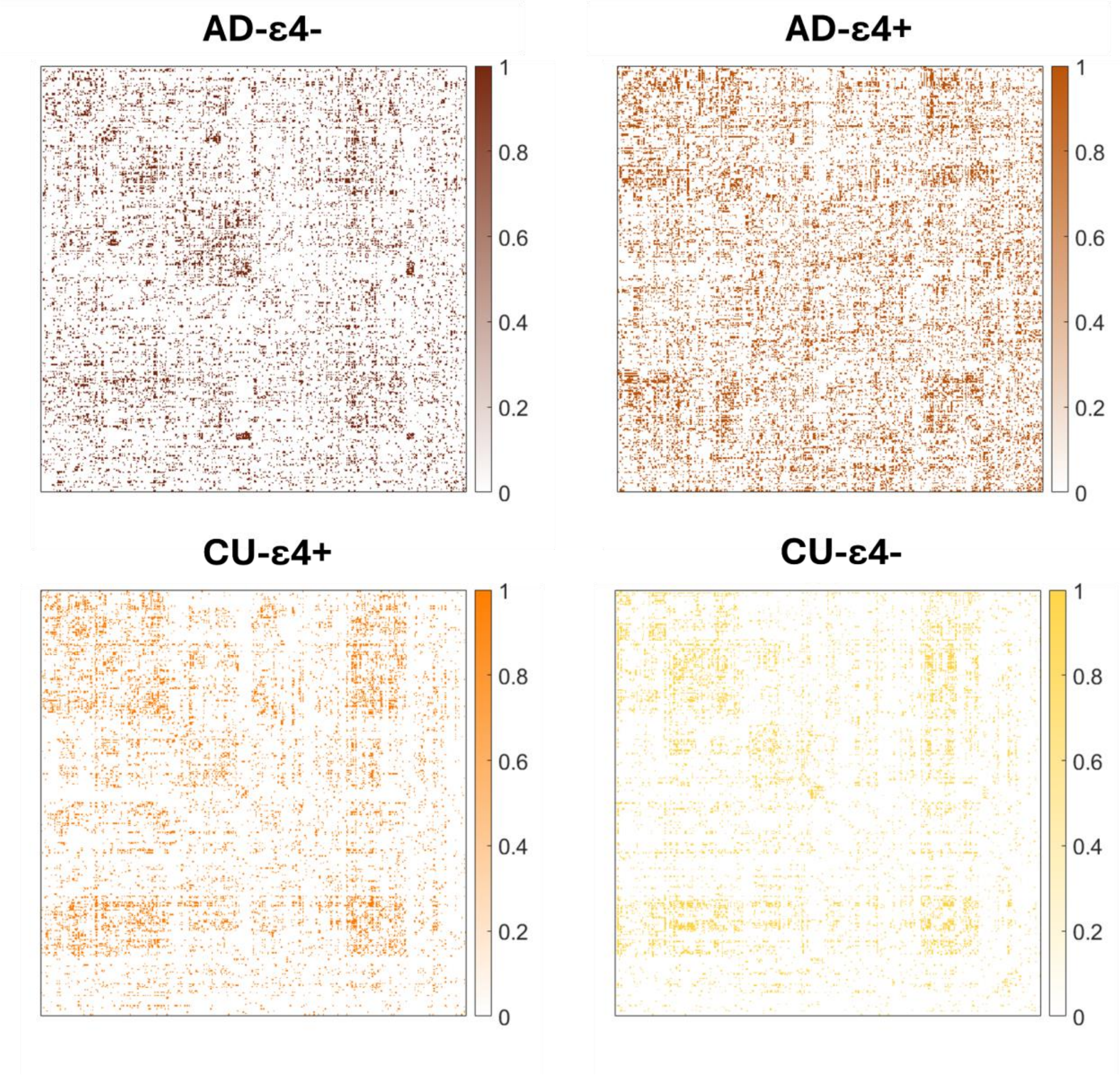


**Figure 5: Binary ICC matrices. Binarized ICCs (>0.6) across groups.**

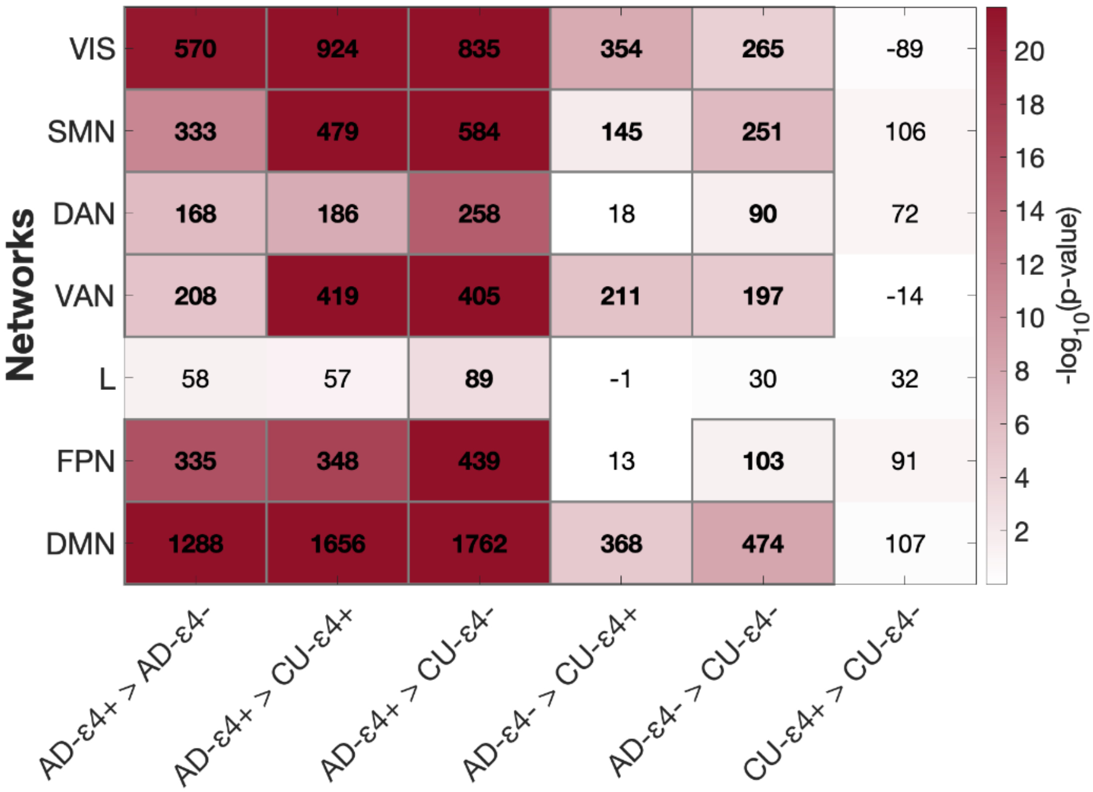


**Figure 6: Results of the ICC statistical analysis.** Heatmap illustrating the corrected p-values obtained from pairwise comparisons between four groups (AD-ε4+, AD-ε4-, ε4+, and ε4-) across seven brain networks. The p-values are transformed using the negative logarithm base 10 (-log10), allowing for a clearer visualization of significance levels. The heatmap is enhanced with effect sizes displayed in each cell, providing additional context to the statistical results. Abbreviations: VIS: Visual, SMN: Somatosensory, DAN: Dorsal Attention, VAN: Ventral Attention, L: Limbic, FPN: Fronto-Parietal, pDMN: Posterior Default Mode Network, aDMN: Anterior Default Mode Network.

#### Fingerprinting results on FC matrices not enriched by INT

In the fingerprinting analysis conducted on raw FC matrices, we found that the SR at which subjects were identified was equal to 100% for all groups. Moreover, group Idiff comparisons ($\chi^2 = 9.38$, $p > 0.05$) did not reveal significant differences across groups; the same result is true for Iself ($\chi^2 = 2.1$, $p > 0.05$). We found significant differences ($\chi^2 = 16.6$, $p < 0.001$) for the Iothers metrics that resulted significantly higher in AD-ε4+ than ε4+ ($z = 9.1$, $p_{FDR} < 0.05$) and ε4- ($z = 11.7$, $p_{FDR} < 0.05$).

13. Orban P, Madjar C, Savard M, et al. Test-retest resting-state fMRI in healthy elderly

persons with a family history of Alzheimer's disease. *Sci Data*. 2015;2:150043. doi:10.1038/sdata.2015.43

22. Reitan RM. Validity of the Trail Making Test as an Indicator of Organic Brain Damage. (84).

23. Amodio P, Wenin H, Del Piccolo F, et al. Variability of trail making test, symbol digit test and line trait test in normal people. A normative study taking into account age-dependent decline and sociobiological variables. *Aging Clin Exp Res*. 2002;14(2):117-131. doi:10.1007/BF03324425

24. Giovagnoli AR, Del Pesce M, Mascheroni S, Simoncelli M, Laiacona M, Capitani E. Trail making test: normative values from 287 normal adult controls. *Ital J Neurol Sci*. 1996;17(4):305-309. doi:10.1007/BF01997792

25. Goldberg A, Rosario I, Power J, Horga G, Wengler K. Strategies for motion- and respiration-robust estimation of fMRI intrinsic neural timescales. *Imaging Neurosci*. 2024;2:1-17. doi:10.1162/imag_a_00326
